# From Normal Variation in Sleep to Clinical Sleep Disorders: Genetic Insights from Over One Million Individuals

**DOI:** 10.1101/2025.11.21.25340447

**Authors:** Lovemore Kunorozva, Jesse Valliere, Chia-Yen Chen, Matthew Maher, Cynthia Tchio, Angus Burns, Yingzhe Zhang, Satu Strausz, FinnGen, Estonian Biobank Research Team, John Winkelman, Daniel J Gottlieb, Tian Ge, Andrew Wood, Michael N Weedon, Samuel E Jones, Susan Redline, Erik Abner, Richa Saxena, Hanna M Ollila, Jacqueline M Lane

**Affiliations:** Brigham and Women’s Hospital Division of Sleep and Circadian Disorders 221 Longwood Avenue, EBRC 101A Boston MA 02115, USA; Broad Institute, Molecular and Population Genetics Program, 415 Main St, Cambridge, MA 02142, USA; Center for Genomic Medicine, Massachusetts General Hospital, Boston, MA, USA; Department of Anesthesia, Critical Care and Pain Medicine, Massachusetts General Hospital, Boston, MA, USA; Biogen, Cambridge, MA 02142, USA; Institute for Molecular Medicine Finland (FIMM), HiLIFE, University of Helsinki, Helsinki, Finland; Functional and Population Genomics, Institute of Genomics, University of Tartu, Tartu, Estonia; Division of Sleep Medicine, Harvard Medical School, Boston, MA, USA; Sleep Disorders Clinical Research Program, Massachusetts General Hospital, Harvard Medical School, Boston, MA, USA; Psychiatric and Neurodevelopmental Genetics Unit, Center for Genomic Medicine, Massachusetts General Hospital, Boston, MA, USA; Center for Precision Psychiatry, Department of Psychiatry, Massachusetts General Hospital, Boston, MA, USA; MRC Human Genetics Unit, Institute of Genetics and Cancer, The University of Edinburgh, Western General Hospital, Crewe Road South, Edinburgh EH4 2XU; University of Exeter Medical School, RILD Building, RD&E Hospital Winford, Barrack Road, Exeter, EX2 5DW, UK; Department of Epidemiology, Harvard T.H. Chan School of Public Health, 677 Huntington Ave, Boston, MA, USA; Medical Service, VA Boston Healthcare System, Boston MA 02132

**Keywords:** sleep, biobank, genetics, genetic association, sleep disorders, medication

## Abstract

Sleep disorders affect over 30% of the U.S population and are linked to increased disease risk and mortality. However, the genetic architecture of sleep disorders and the overlap of sleep disorders with habitual sleep quality, quantity, and timing have been poorly characterized. In addition, it is unknown if clinical sleep disorders are extremes of habitual sleep traits. Here, we systematically investigated the genetic basis of seven clinical sleep disorder traits, and sixteen medications used for sleep problems in 1,600,000 individuals. We identified 590 genetic associations for sleep apnea, insomnia, restless legs syndrome, narcolepsy and a combined sleep disorder phenotype, of which 367 were previously unreported. Additionally, we discovered 142 genetic associations with sleep medication use. While overall genetic architecture was shared across sleep traits, we found unique genetic factors for the different sleep disorders that reflect fundamentally different biological mechanisms including for example autoimmune processes with narcolepsy and skeletal morphology in sleep apnea. Furthermore, sleep genetic factors showed a broad multi-omic impact on gene and protein expression levels. These findings suggest that while clinical sleep disorders share genetic architecture with each other and with variation in sleep patterns within the general population, they are not simply extremes of normal sleep variation but involve unique biological mechanisms. These results advance our understanding of sleep disorders and suggest potential novel therapeutic targets.

## INTRODUCTION

Clinically significant sleep disorders are highly prevalent, affecting an estimated 25-30% of adults worldwide.^1–3^ Of 80 known sleep disorders^4^, the most common are insomnia (30%)^5,6^, sleep apnea with prevalence ranging from 17-34%^7^, restless legs syndrome (RLS) 4-14%^8^, and narcolepsy 0.02-0.04%.^9^ The impact of sleep disorders on the individual includes insufficient or incorrectly timed sleep, excessive daytime sleepiness and functional impairment, all of which can have debilitating effects on health, particularly increased risk for psychiatric^10^, cardiometabolic disorders,^11,12^ and other chronic conditions such as cancer and neurodegeneration. Thus, understanding the causes of sleep and circadian disorders and the mechanisms underlying their health consequences could improve disease management, alleviating the burden at the level of individuals and families, and at the societal, medical system, and economic level.

Sleep and circadian rhythms affect the timing and duration of sleep, and the quality of both sleep and wakefulness. Sleep traits are influenced by both internal (for example genetic and hormonal)^13–15^ and external factors (for example environment, cultural factors, comorbidities or drugs).^16,17^ However, the distinction between sleep symptoms and clinically diagnosed sleep disorders often reflects not only underlying biology but also symptom severity and access to health care. Additionally, the connection between normal regulation of sleep and sleep dysregulation in clinical sleep disorders is poorly understood. As circadian and sleep phenotypes have a heritable and an environmental component, genetics provides an opportunity to advance our understanding of the biological pathways and molecular mechanisms underlying sleep and circadian regulation, sleep disorders, and their links with other diseases and response to environmental risk factors.

Genome-wide association studies (GWAS) have successfully identified genetic variants associated with several sleep disorders and have provided new insights into their biological underpinnings.^18^ Furthermore, sleep medication use is an informative proxy that captures underlying clinically meaningful sleep problems. Consequently, genetics of sleep medication use can be an additional tool to gain insight into the biological processes that drive individual variability in sleep architecture, circadian timing, and sleep disorder and related disease susceptibility. Although sleep is often considered primarily a neurological function, mounting evidence shows that it is modulated by diverse biological systems, including metabolic and immune pathways.^19,20^ Examples include the strong influence of BMI on sleep apnea risk^21^, autoimmune associations in narcolepsy^22^, and iron deficiency as a contributing factor in RLS.^23^

Furthermore, sleep disorders with overlapping symptoms may have distinct biological mechanisms. For instance, both sleep apnea and narcolepsy result in excessive daytime sleepiness, yet they differ markedly in etiology and treatment. Sleep apnea symptoms arise from airway obstruction and consequent sleep fragmentation and gas exchange abnormalities and is generally treated with continuous positive airway pressure (CPAP) or other anatomic therapies, whereas narcolepsy is an autoimmune disease generally treated with stimulants.^20,24^ Understanding the physiological and molecular relationships between symptoms and underlying disorders is essential to improving diagnostic accuracy and guiding appropriate therapeutic interventions.

To advance the understanding of sleep traits, we conducted a meta-analysis of GWAS of sleep disorders and medications and compared the genetic architecture across these traits and to eight self-reported^25–30^ and eight actigraphy derived objectively measured sleep and circadian traits^31^ in up to five cohorts and biobanks in 1.6M individuals (**Fig. 1, Table S1-2)**.

**Figure 1.**
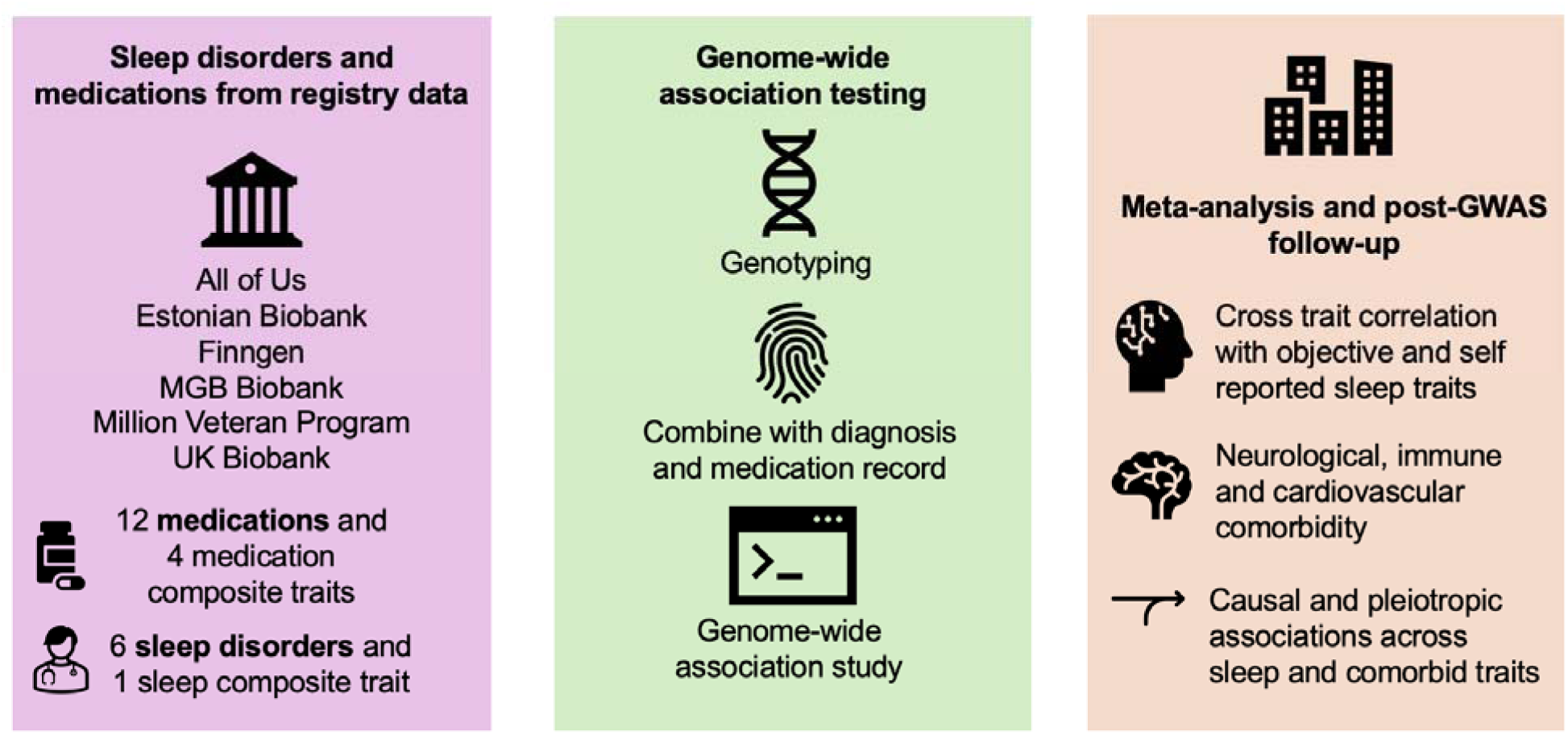
Overview of the analytical strategy for examining the genetics of sleep disorders. We performed genome-wide association testing and meta-analysis across six cohorts including five discovery cohorts and replication in the Estonian Biobank for altogether 23 traits. We then performed cross-trait analysis, genetic correlation, and examined robustness of signals across cohorts and followed up individual genetic variants.

## RESULTS

### Genetic analysis discovers novel loci across clinical sleep disorders and sleep medications

We analyzed genetic associations across seven clinical sleep traits (CSD) and sixteen traits for medications commonly prescribed for insomnia and related sleep complaints (**Fig. 1, Table S1-3**). We meta-analyzed genome-wide association data from participants of European ancestry from five cohort comprising UK Biobank (UKB), Mass General Brigham Biobank (MGBB), All of Us, Million Veteran Program and FinnGen (N = 1,630,075 individuals, **Tables S1-3**).

Through LD pruning and fine-mapping, we discovered 590 significant independent genetic associations for sleep disorders (**Table S3, Fig 2**). These include 357 variants associated with sleep apnea (314 previously unreported, N=312,402 cases), 35 variants with clinically diagnosed insomnia (10 previously unreported, N=178,316 cases), 122 variants with RLS (41 previously unreported, N=28,814 cases), and 2 loci with narcolepsy (four variants at HLA and one previously unreported association outside HLA, N=2,172 cases). Additionally, we observed 142 significant independent variants for medication use in the meta-analysis of UKB, MGBB, and in FinnGen (**Table S3, Fig 2**). Overall, the results indicated a robust polygenic signal that aligned with expected λ_GC_ distribution across traits (**Table S4**). SNP-based heritability estimates ranged from 2.6% to 14.1% (**Table S4, Fig S2**) and were consistent with earlier studies.^32,33^

**Figure 2:**
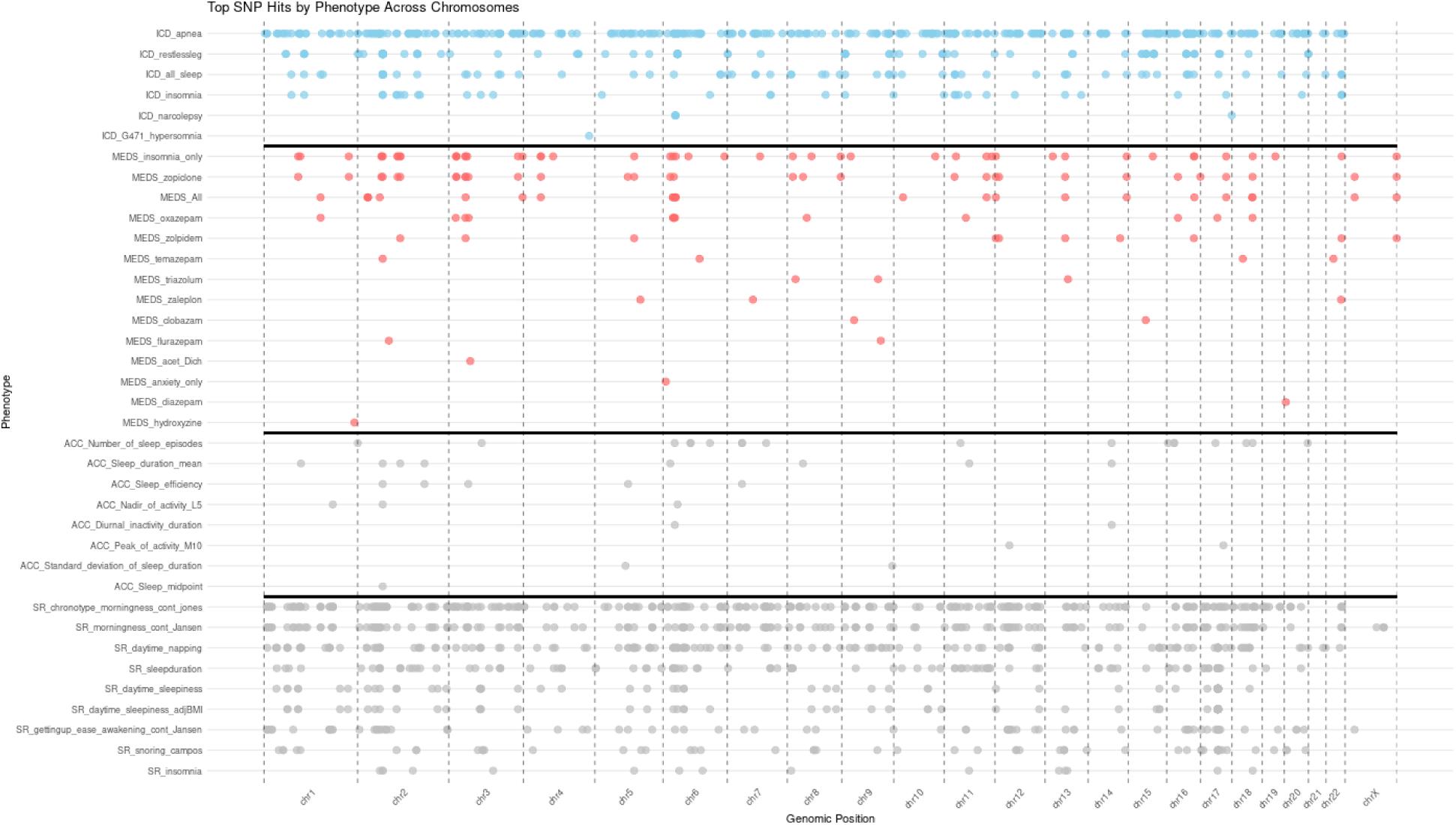
Genomic distribution of Top SNP hits across clinically diagnosed conditions, medication usage, self-reported traits, and actigraphy-derived measures grouped by chromosome (x-axis) and categorized by phenotype (y-axis). Each dot represents a significant SNP, color-coded by phenotype group, with the density and clustering of SNPs highlighting potential genomic hotspots. Chromosomes 1, 6, 11, and 17 emerge as key loci with pleiotropic effects, influencing multiple phenotypes.

We then replicated the findings in the Estonian biobank (N = 205,822 individuals) where power analysis indicated expected nominal replication rates (α = 0.05) of approximately 10-20% for sleep apnea, 5-15% for clinical insomnia and 25-35% for RLS. The observed nominal replications were 61/357 associations (17.1%) associated with sleep apnea (N = 12,000 cases), 4/35 associations (11.4%) for clinically diagnosed insomnia (N = 12,962 cases), and 36/122 associations (29.5%) with RLS (N = 2,180 cases, **Table S5**), and consistent with these expectations given limited power for smaller effects. Our results showed a number of consistent associations with earlier reported CSDs such as the *MEIS1* association with RLS, *PAX8* locus with insomnia, *FTO* locus with sleep apnea, and a remarkably strong association at the HLA locus with narcolepsy providing a proof of principle of robust and reproducible associations across cohorts (**Fig 2, Table S3, S5**).^34–36^

### Gene enrichment and Pathway analysis of sleep disorders

Genetic associations with insomnia were localized in genes upregulated in brain tissues, particularly in anterior cingulate cortex - a key region in insomnia physiology and genes downregulated in adipose tissue from GTExv8. Additionally, we identified 33 transcription factors that were enriched in the clinical insomnia associations. Finally, cell type specific expression enrichment supported enriched expression in midbrain neurotypes and fetal heart (**Fig. S3**). In addition, we observed 26 transcription factor targets in RLS (**Fig. S4**), and enrichment of gene ontology categories for locomotion and neuronal differentiation, which were supported by single cell type association in midbrain neurons and kidney cell types in agreement with a known neurobiological origin, and higher comorbidity of RLS in individuals with kidney disease^37^ (**Fig S4**). Finally, analysis of sleep apnea identified a clear neuronal signal across several brain regions including hypothalamus, hippocampus and cortex but additionally a signal with vasculature and esophagus highlighting both central nervous system and peripheral tissues in the multifactorial mechanisms underlying sleep apnea (**Figure S5**). Additionally, the analysis identified 143 transcription factor targets (**Table S6**), and gene ontology biological processes of neurodevelopment, respiratory gas exchange and growth and morphogenesis which all were also supported by single variant associations (**Table S6**).

Specifically, reduced BDNF signaling has been repeatedly linked to poor sleep quality, reduced slow-wave activity, and vulnerability to mood and anxiety traits.^38,39^ Additionally, we observed insomnia associations with *NKAIN2* ATP-dependent sodium-potassium transport channel that has previously been associated with mood traits^40^, and *CNTNAP5* that belongs to neurexin family and is involved in cell-cell adhesion and neuronal connectivity (**Fig 2, Table S3, S7**). Furthermore, we observed new signals for known RLS loci (*BTBD9*) but where the lead variants were not in LD with the lead variants from earlier studies.^38^ Furthermore, we found associations at previously unreported loci, including *ZFHX3* that has been implicated in retinal sensitivity and circadian responses to light.^41^ Other previously unreported associations included *NCAPD3* and *IQCH,* that connect RLS with energy metabolism and cardiovascular disorders that are often comorbid with RLS.^42^

The largest number of variants were observed with sleep apnea where we observed 314 previously unreported associations (**Table S3, S7**). In addition to loci related to cardiometabolic traits, we discovered associations at *ARNTL* locus, a core circadian regulator.^43^ Additionally, the associations included *IGLON5* that encodes a neural adhesion protein. A rare autoimmune sleep disorder anti-IGLON5 disease manifests with antibodies towards the IGLON5 protein and causes several sleep problems, including sleep apnea.^44^

Additionally, we observed several developmental and craniofacial associations. For example, we saw associations with *SOX6*, a transcription factor implicated in skeletal development and craniofacial morphology relevant to airway patency.^45^ In addition, we observed bone morphogenesis protein 5 (*BMP5)* that induces bone and cartilage development^46^ and an association at the *COL11A2* locus that encodes a type XI collagen component. The associations with circadian rhythm, skeletal, bone, and collagen genes suggest that sleep apnea arises from multiple parallel biological mechanisms, involving both central regulation of breathing and structural factors that affect upper airway patency, consistent with findings from clinical studies ^47,48^ (**Fig 2, Table S3**).

### Sleep medications identify previously unreported associations for sleep

Sleep problems are also routinely treated by sleep medications when the primary indication for treatment is not necessarily a sleep disorder. We observed 142 associations including at known loci for sleep disorders (e.g. *MEIS1, CACNA1C, OLFM4*, *PAX8* and *TCF4,* **Table S3**). Approximately half (80/142, 56%) of the loci that were associated with medications were not associated with clinical sleep traits at a genome-wide significant level but revealed additional genes important in the regulation of sleep, circadian rhythms, metabolism, and stress-related processes. We identified neuronal and synaptic associations including adrenergic and gabaergic signaling at *ADRA1D*, *DCC*, *ECRG4, FMN2*, *GABRA4*, *GABBR1* and *TAOK2* highlighting the strong neurobiological contribution to sleep medication use.

### Genetic associations implicate pleiotropy across sleep disorders

We next estimated the pleiotropy between sleep disorders defining a variant as pleiotropic if it reached genome-wide significance (p < 5 × 10□□) for that trait and simultaneously showed genome-wide significant associations with at least one additional trait. After applying this criterion and performing LD-pruning to ensure independence, we discovered that 14 (40%) of the 35 insomnia associated variants were pleiotropic with the other CSDs and 18 (51%) also associated with medications (**Table S7-9**). Furthermore, 63 (18%) of 357 sleep apnea variants were pleiotropic and associated with other CSDs and 30 (8%) with medications. Finally, 15 of the 122 (12%) RLS-associated variants were pleiotropic with other CSDs and 54 (44%) with medications (**Table S7)**. Given the pleiotropic associations, we wanted to understand if individual genetic variants cluster by trait category.

Sleep apnea and insomnia variants clustered with sleep fragmentation from objectively measured accelerometer traits (**Fig 3a-b**), which broadly capture sleep quality, timing and variability, recapitulating the shared genetic architecture of sleep fragmentation as a phenotype in both traits. Surprisingly, a subset of variants that were associated with increased sleep apnea risk showed an association with decreased number of sleep episodes during the night with strong and significant opposite effect estimates (**Fig 3a**). Such associations might reflect different mechanisms underlying OSA pathophysiology, with high sleep fragmentation variants possibly reflecting low arousal threshold while low fragmentation variants may reflect high arousal threshold and allow expression of other endotypes such as low neuromuscular compensation. This bidirectional association was seen in variants such as *APOE*, which in our study increased the risk for sleep apnea, while lowering the risk for sleep fragmentation or increasing sleep need. A subset of variants that were associated with increased insomnia risk also showed an association with decreased number of sleep episodes during the night and may reflect increased sleep need (**Fig 3a**). Furthermore, we observed that the variants associated with clinical insomnia were associated with sleep medication use and clustered together by effect size with sleep medications (**Fig 3c**) showing that sleep medications used for insomnia might identify shared genetic signatures.

**Figure 3.**
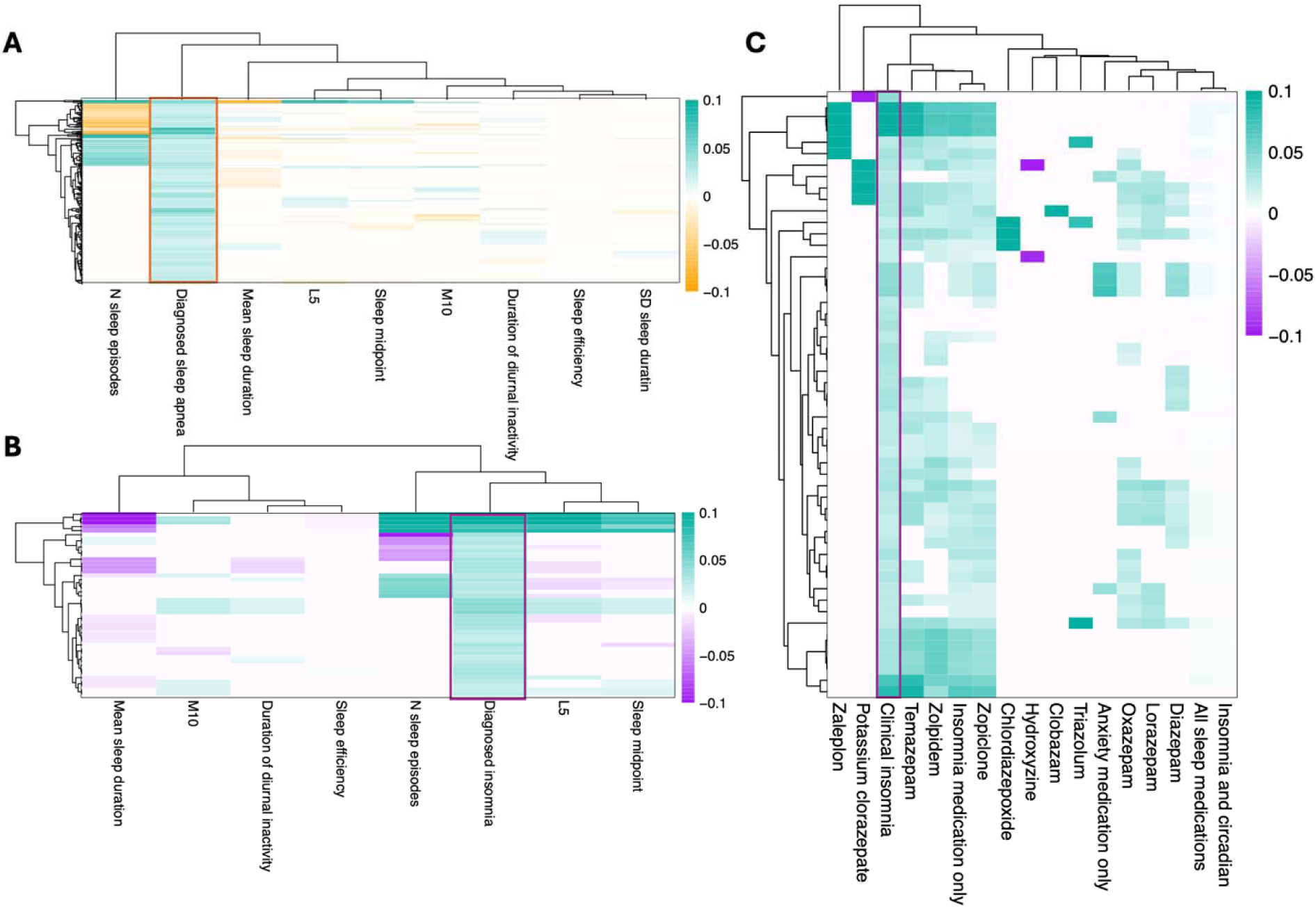
Clustering of genome-wide significant associations for sleep apnea and insomnia based on the effect sizes across accelerometer-derived sleep traits and sleep medication use. Using all genome-wide significant lead variants for either sleep apnea or clinical insomnia we show the clustering of effect size (beta) with other sleep traits at single variant level. A) GWAS associations of sleep apnea variants with accelerometer traits. B) GWAS associations of insomnia lead variants with accelerometer derived traits. C) GWAS associations of insomnia associations with sleep medications.

### Bayesian clustering highlights distinct mechanisms in sleep disorders

The connection between sleep apnea and obesity is well known. However, as suggested by novel variant at craniofacial gene loci the analysis also identified non-BMI related associations. To assess which sleep apnea variants were mediating liability to sleep apnea through BMI, we performed Bayesian clustering of effect sizes. We observed that 18 contributed primarily to BMI (PP > 0.8), 98 were shared between sleep apnea and BMI whereas 69 variants had posterior probability over 0.8 for contributing only to sleep apnea (**Figure 4a**). Furthermore, the craniofacial and bone related genes had high posterior probability for associating only for sleep apnea (*BMP5*, PP = 0.99; *COL11A2*, PP = 0.98).

**Figure 4.**
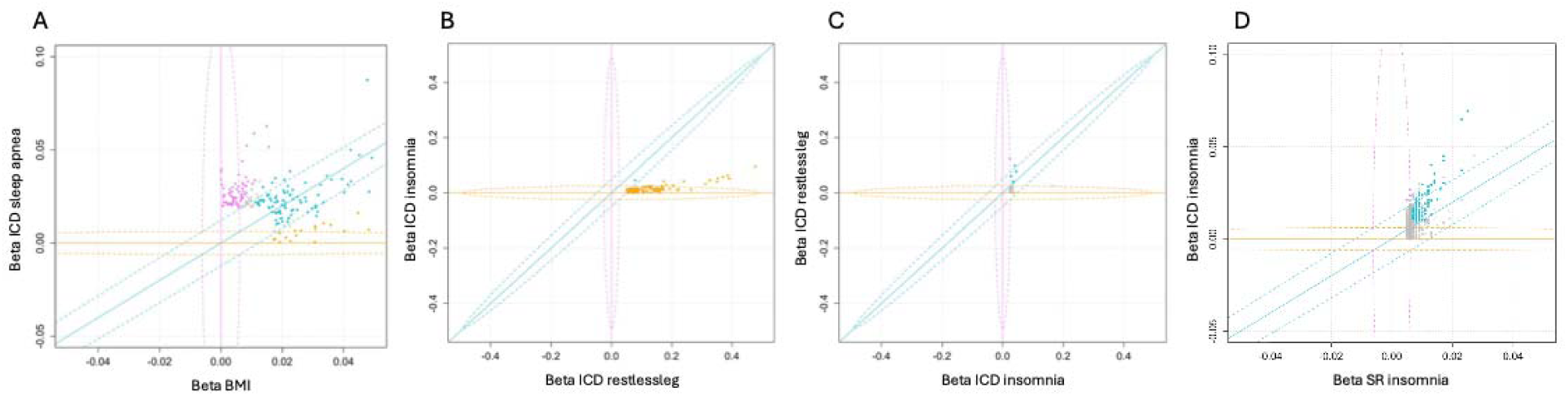
Bayesian clustering of individual variants shows unique and shared signals across sleep traits. **A)** Comparison of effect sizes of sleep apnea associating variants suggest a subset specific for BMI (yellow), shared variants (cyan) and variants specific for sleep apnea (pink). **B)** Comparison of effect sizes between RLS variants in RLS vs. insomnia shows that the majority of RLS associations are specific for RLS **C)** Insomnia associations compared between clinical insomnia and RLS show clustering of either clinical insomnia, RLS or shared associations, **D)** Self-reported insomnia associations have lower effect size in self-reported insomnia than in clinical insomnia.

Furthermore, a longstanding question in the field of RLS and insomnia has been how much of the genetic signal between RLS and insomnia is shared. To test this, we performed Bayesian clustering of effect sizes between RLS and clinical insomnia (**Fig 4b-c**). We observed that all variants that were discovered associating with RLS showed high posterior probability (>0.8) of being uniquely associated with RLS, with one signal at *EXD3* locus shared between clinical insomnia and RLS (PP = 1). Furthermore, clinical insomnia associations highlighted two loci that were primarily associated with RLS, *MEIS1* consistent with earlier studies^30,49,50^, and *PRMT6,* as well as five additional shared loci and one locus specific to clinical insomnia, *PAX8* (PP = 0.8).

It is possible that habitual sleep traits and CSDs share underlying genetic variants, but that variant effects are larger in clinical disorders than in habitual sleep traits. Using effect size clustering, we observed that variants shared between clinical insomnia and reported insomnia symptoms had higher effect sizes for clinically diagnosed insomnia than insomnia symptoms, with a subset of variants appearing specific to clinical rather than insomnia complaints (**Fig 4d**). Additionally, we observed that effect size estimates for variants that associated with sleep apnea were higher than those for snoring, including the variants that were discovered in genome-wide association study of snoring (**Table S7, S10**). This may reflect a more severe phenotype in clinical versus self-reported traits, lower accuracy in self-reported symptoms or fundamental underlying phenotypic differences not captured by clinical coding.

### Genetic correlations between traits indicate shared etiology between medications and CSDs

We next examined genome-wide genetic correlation and polygenic risk score associations between the main CSDs and neuropsychiatric traits where we saw a substantial shared genetic architecture of insomnia, RLS and sleep apnea with neuropsychiatric traits (**Fig. S6, Table S11a-b**). Furthermore, we observed that CSDs and commonly prescribed insomnia medications showed significant and strong genetic correlation with clinical insomnia and RLS (rg > 0.4 P < 2*10^-55^, **Fig 5a, b)**. Furthermore, we observed strong genetic correlations between CSDs, and habitual sleep phenotypes derived from actigraphy and self-reported sleep traits (**Fig 5a, c**). All sleep disorders and particularly clinical insomnia were strongly associated with lower sleep efficiency and delayed sleep timing, reinforcing the connection between insomnia and non-restorative, poor sleep. Sleep apnea, a sleep disorder characterized by excessive daytime sleepiness, and fragmented sleep, showed genetic correlations with accelerometer derived measures of disrupted sleep, specifically sleep fragmentation and its component trait, number of sleep episodes, suggesting a common genetic basis for impaired sleep continuity and recurrent awakenings. Furthermore, narcolepsy, sleep apnea and hypersomnia all correlated with daytime napping, daytime sleepiness and number of sleep episodes and diurnal inactivity highlighting the symptomatology of daytime sleepiness across these traits at genetic level (**Fig 5, Table S11**).

**Figure 5.**
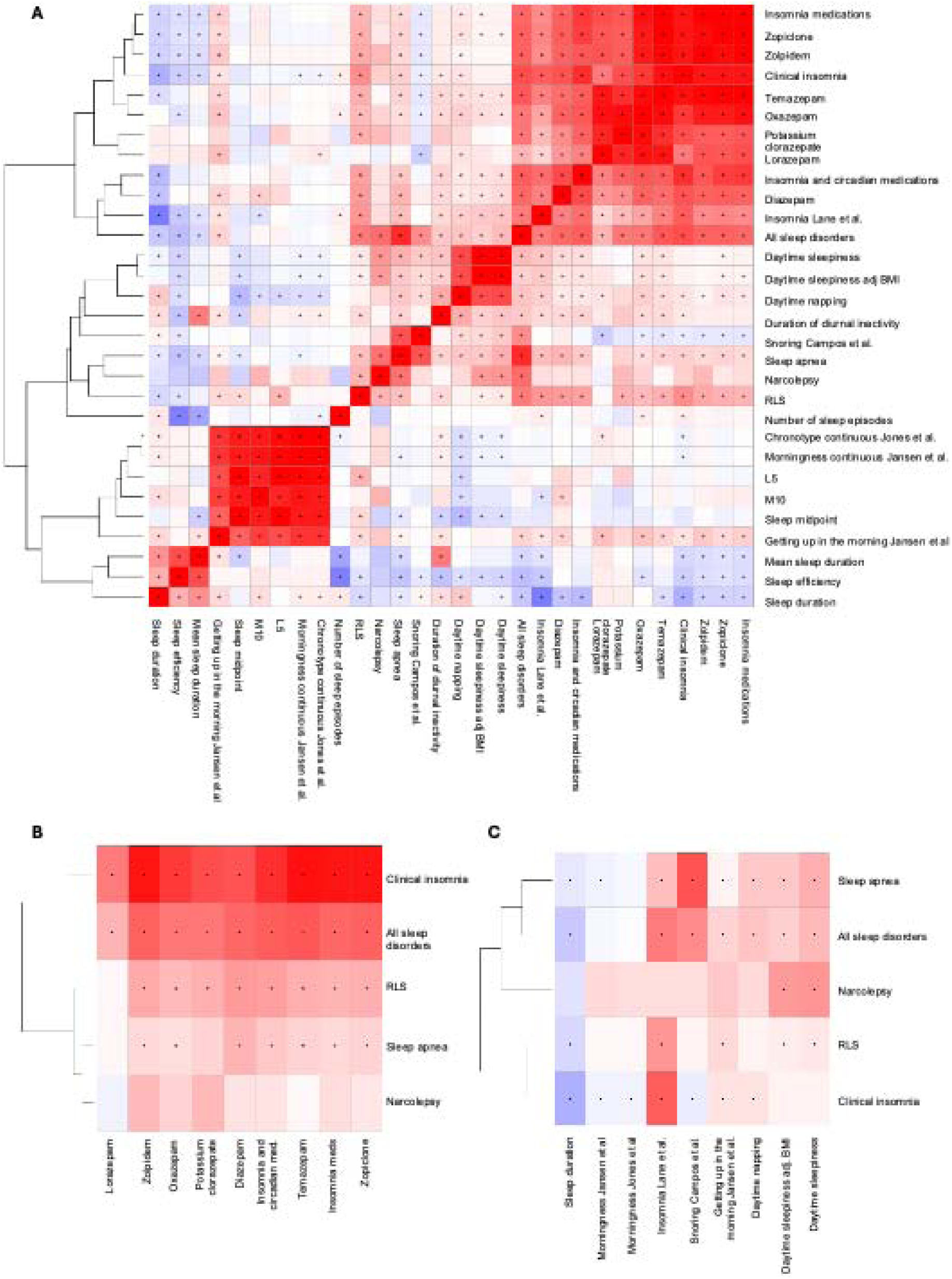
Genetic correlations between clinical sleep disorders, commonly used sleep medications, habitual sleep traits and objectively measured actigraphy traits. **A)** Genetic correlation across all traits. **B)** Genetic correlation between sleep disorders by clinical diagnoses and medications. **C)** Genetic correlation between clinical diagnoses and self-reported traits. Correlation values range from −1 (blue, negative correlation) to +1 (red, positive correlation), with the intensity of color representing the strength of the correlation. The dendrogram (left) clusters sleep disorders based on genetic correlation patterns, offering insights into shared biological pathways and therapeutic overlap.

In addition, RLS showed negative correlations with sleep efficiency and positive associations with fragmented sleep episodes, supporting its characterization as a disorder of disrupted sleep. Collectively, these findings highlight substantial genetic overlap among pharmacologic treatments, clinical sleep disorders, and habitual sleep traits (**Fig 5, Table S11**).

### Genetic variants for sleep disorder and medication use associate with systemic diseases in PheWAS

While sleep disorders may result in similar phenotypic symptoms, separate triggers and risk factors give rise to each sleep disorder. To explore the association with systemic diseases, we performed a PheWAS of all genome-wide significant loci identified for CSD and medications. Loci for sleep traits and medication use associated with neuropsychiatric, cardiometabolic and immune traits, in agreement with earlier epidemiological observations and recent genetic studies (**Table S8a-l**).^38,51,52^ The PheWAS also identified the multi-omic impact of sleep trait loci on gene expression, gene splicing and circulating protein levels, results which will help in the prioritization of effector genes at each locus. Additionally, we tested previously implicated risk factors specifically including low ferritin level as a risk factor for RLS,^38,53^ and high BMI for sleep apnea^11,54^ using Mendelian randomization. We observed a significant association from low ferritin to high RLS (**Fig. S7a, Table S12**) with no effect on insomnia (**Fig. S7b**). We also validated the earlier reported association between an increase in BMI and the increased risk for sleep apnea (**Fig. S7c**).

## Discussion

Here, we systematically investigated the genetic architecture underlying sleep disorders, habitual variation in sleep, objective sleep traits, and sleep-related medication use. Our meta-analysis identified 590 genome-wide associations for CSDs (367 previously unreported) and 142 associations linked to sleep medication use. We identified novel biologically relevant associations for sleep traits including craniofacial genes in sleep apnea, neuronal signaling molecules in medication use and expand the knowledge of cardio-metabolic and neurodevelopmental genetic links to clinical insomnia and RLS. Additionally, our findings also implicate shared genetic architecture in sleep traits.

Although diagnosis and questionnaire-based assessments remain standard in genetic epidemiology, emerging evidence from other fields suggests that analyzing medication use offers complementary insights into disease biology, particularly for underdiagnosed diseases with a large population burden, such as sleep disorders. Our results support this, revealing that a subset of variants associated with insomnia and other sleep disorders also predict sleep medication use, but that about half of the variants associated with sleep medication use reveal additional neurobiological associations not seen for sleep disorders. Thus the association between insomnia and medication use may arise from insomnia symptoms that are mild or subclinical and are perhaps treated without a specific clinical diagnosis of insomnia.

By identifying 590 genome-wide significant associations across CSDs including 357 for sleep apnea, 35 for insomnia, 122 for RLS, and 5 for narcolepsy our study demonstrates the power of large-scale, multi-cohort GWAS to refine the genetic foundations of sleep phenotypes. In doing so, we both validate previously reported loci and uncover novel associations, offering fresh insights into shared and disorder-specific pathways.

Our findings highlight a substantial overlap between genetic architecture of CSDs and habitual sleep traits. For example, the high number of shared variants between clinical sleep apnea and self-reported snoring reinforce the concept that part of sleep disorders can be traced with relatively simple questions about sleep characteristics such as snoring. *MEIS1* similarly exhibited consistent associations across RLS, clinical insomnia, and self-reported sleep symptoms, underscoring its pleiotropic effects across poor nighttime sleep. Genes such as *CACNA1C* and *MEIS1* were also linked to psychiatric traits, suggesting that common neurobiological pathways influence sleep and mental health. Additionally, these findings may suggest heterogeneity of insomnia endotypes. For example, RLS is related to sleep-onset insomnia. However, the genetic architecture of anxiety and circadian triggered insomnia are largely different. Consequently, genetic architecture might help to elucidate and thereby guide therapy or development of therapeutic targets.

Finally, despite the advances made in this study, several limitations warrant attention. Genetic analyses of diagnostic codes and medications from biobank-scale cohorts used phenotypes derived from electronic health records, which may have limited sensitivity and specificity.^55,56^ Country-specific diagnostic practices and medication prescription patterns are additional limitations. Furthermore, the extent of comorbidities varied across cohorts, as we included data from large-scale cohorts and biobanks, a hospital-based biobank, and from US veterans. Most cohorts included were of European ancestry, which limits the generalizability of findings to other populations.^57^ Increasing representation of diverse populations in genetic studies is essential to identify ancestry-specific loci and ensure equitable application of results. Future efforts should integrate multi-omics data, including transcriptomics and proteomics, to refine mechanistic understanding of sleep regulation. Pharmacogenetic studies, especially in diverse populations, could improve individualized treatment approaches.^58^ Addressing the heterogeneity of sleep disorders, particularly subtypes of insomnia, will be important for developing targeted therapies. Integrating genetic, environmental, and behavioral data could enable more precise models of sleep biology and accelerate progress toward personalized medicine in sleep health.

Overall, our findings implicate unique genetic architecture for distinct sleep disorders with neuronal, immune, developmental and cardiovascular components. Additionally, sleep medications provide a complementary and novel angle for understanding sleep problems potentially through capturing genetic variation that is not covered with questionnaire or diagnosis codes. Our findings show a robust polygenic signal, large disease specific components and strong connection with comorbid traits across sleep and sleep medication traits.

## METHODS

### COHORTS

#### UK biobank

The UK Biobank (UKB) is a population-based study containing over 500,000 individuals of mainly European ancestry.^59^ The participants were recruited to the study between 2006 and 2010, were aged between 37 and 73 years of age and were residents of the United Kingdom. The study is a combination of different lifestyle measures, genotypes, electronic health record data, 7-day accelerometry and questionnaire data, and the health record data is updated frequently to capture the health trajectories of participating individuals.

##### UKB Phenotypes

In the UK Biobank, we extracted health data from primary care records. General Practitioner (GP) data was available for approximately 230,000 participants. Additionally, we used in-patient records (Hospital Episode Statistics) mapping to ICD codes **(Table S1)**. Any sleep disorder cases were identified as having any of the diagnoses (**Table S1**). We extracted medication records on sleep-related prescriptions for insomnia, insomnia and anxiety, and circadian medications (**Table S1**). Any sleep medication and any sleep specific medication, all drugs prescribed for sleep regardless of indication, were measured as binary variables.

##### UK Biobank Genetic analyses

The UKB genotyped 488,477 participants: 49,950 on the Affymetrix (Thermo Fisher Scientific) UK BiLEVE Axiom Array and 438,427 on the highly similar Affymetrix UK Biobank Axiom Array. These arrays captured up to 825,927 SNPs and short indels, with variants prioritized for known coding variants, those previously associated with disease and ancestry-specific markers that provide a good imputation backbone. DNA was extracted from blood samples taken at baseline visit, between 2006 and 2010, and genotyping was carried out in 106 sequential batches, giving genotype calls for 812,428 unique variants in 489,212 participants. After removing high missingness and very rare variants, as well as poor-quality samples, these genotypes were phased using SHAPEIT3 and imputed to the Haplotype Reference Consortium (HRC) and to a merged UK10K and 1000 Genomes phase 3 reference panel^59^, both in genome assembly GRCh37 using IMPUTE2. This resulted in 93,095,623 autosomal SNPs, short indels and large structural variants in 487,442 individuals and 3,963,705 markers on the X chromosome. For more details, see Bycroft et al.^59^ Genetic association testing was done using Regenie^60^

##### Mass General Brigham biobank

The MassGeneralBrigham (MGB) Biobank is a hospital-based cohort study from the MGB healthcare network with electronic health record (EHR), genetic, and lifestyle data.^61^ The MGB biobank participant recruitment and population cohort has been described previously.^61^ We used up to 51,053 individuals (24,672 cases, 26,318 controls) of European genetic ancestry from the full MGB biobank who had genetic data available.

##### MGB Biobank Phenotypes

For the MGB cohort, ICD-9 and ICD-10 diagnostic codes were obtained from the Research Patient Data Registry (RPDR), a centralized clinical data repository that integrates information across multiple hospitals in the MGB network.^62^ RPDR aggregates structured clinical data including inpatient and outpatient diagnoses, laboratory results, procedures, and medication records. For this study, we extracted diagnostic information for participants linked to the MGB Biobank and identified sleep-related phenotypes using ICD codes (**Table S1**).

We extracted prescription data related to sleep and circadian disorders, including insomnia and anxiety, from the MGB-RPDR using RxNorm Concept Unique Identifiers (RxCUIs). All medications previously identified in the UK Biobank for these conditions were systematically searched and mapped within the MGB-RPDR to ensure consistency and facilitate cross-cohort harmonization.^63^ To account for regional differences in prescribing practices, we consulted a local practicing physician to identify U.S. medications that are either unavailable or infrequently used in the UK and to determine equivalent drug names where applicable. Medications from the MGB Biobank were then mapped to their UK Biobank counterparts using RxCUIs to support cross-cohort comparison and meta-analysis. Most insomnia and anxiety medications in the U.S. cohort, such as chlordiazepoxide, diazepam, hydroxyzine, lorazepam, oxazepam, temazepam, zaleplon, zolpidem, clobazam, triazolam, and clorazepate dipotassium had direct or closely aligned equivalents in the UKB. Zopiclone, which is not approved in the U.S., was mapped to its pharmacologic analogue, eszopiclone. We included medications with at least 100 recorded prescriptions in the MGB Biobank cohort (**Table S1**).

#### Genotyping and association testing

The MGB Biobank genotyped 53,297 participants on the Illumina Global Screening Array (’GSA’) and 11,864 on Illumina Multi-Ethnic Global Array ("MEG"). The GSA arrays captured approximately 652K SNPs and short indels, while the MEG arrays captured approximately 1.38M SNPs and short indels. These genotypes were filtered for high missingness (>2%) and variants out of HWE (P < 1 × 10^−12^), as well as variants with an AF discordant (P < 1 × 10^−150^) from a synthesized AF calculated from GnomAD subpopulation frequencies and a genome wide GnomAD model fit of the entire cohort. This resulted in approximately 620K variants for GSA and 1.15M for MEG. The two sets of genotypes were then separately phased and imputed on the TOPMed imputation server (Minimac4 algorithm) using the TOPMed r2 reference panel. The resultant imputation sets were both filtered at an R2 > 0.4 and a MAF >0.001 (minor allele frequency), and then the two sets were merged/intersected resulting in approximately 19.5M GRCh38 autosomal variants. The sample set for analysis here was then restricted to just those classified as EUR (*N* = 54,452) according to a metric of being +/− 2 SDs of the average EUR sample’s principal components 1 to 4 in the HGDP reference panel. Association testing was done with REGENIE^64^ version 3.2.8 adjusting for age and sex population structure PC1-10 and genotyping chip.

#### FinnGen

FinnGen is a public-private cohort established in 2017.^65^ The study combines genetic data with EHR data, including International Classification of Diseases (ICD) codes, derived from primary and specialized care registers, hospital inpatient and outpatient visits and drug prescriptions of 500,348 participants (release 12). We used up to 500,348 Finnish individuals (**Table S1**) of European genetic ancestry from FinnGen R12 that had genetic data available. The Finnish population has been previously described.^66^

##### FinnGen phenotypes

We utilized the ICD-coded registry data available through Finnish national health registers, linked to genomic data for participants included in FinnGen Release 12 (Table S1). All ICD codes were obtained via standardized phenotype mapping protocols developed by the FinnGen consortium to support large-scale genetic association studies (FinnGen 2024). To ensure consistency in phenotype definitions and support cross-cohort comparison, the ICD-10 codes used to define sleep-related phenotypes in the UK Biobank were systematically mapped within FinnGen’s curated registry data^63^

We extracted prescription data for medications related to sleep and circadian disorders, anxiety, and insomnia using Anatomical Therapeutic Chemical (ATC) classification codes from both primary and secondary care records. Medication phenotyping was based on ATC codes as curated by the FinnGen project (FinnGen, 2024), and focused on agents commonly prescribed for sedative, anxiolytic, or circadian-modulating effects (**Table S9**) with at least 100 recorded cases.

##### FinnGen genetic analysis

Specifically, Kurki et al.^66^ defined a set of individuals of Finnish genetic ancestry, by performing a PCA using a subset of variants with a high-quality genotype probability and low missingness. These genotypes met the following criteria: an IMPUTE2 genotype information score ≥0.95, missingness ≤0.01 (i.e., according to genotype probability) and MAF ≥0.05. Because association analyses are performed using the LMM method in Regenie adjusting for age, sex, principal components, genotyping platform, and study specific additional covariates (see Kurki et al., for details).

##### The Million Veteran Program

The Million Veteran Program (MVP) is a longitudinal cohort study of diverse U.S. Veterans exmaining how genes, lifestyle, military experiences, and exposures affect health and wellness.^67,68^ It combines genetic data with EHRs of 635,969 participants (data freeze 4) across four ethnic groups. In this study we extracted data from the publicly released summary statistics for MVP.

##### The Million Veteran Program Phenotypes

For this analysis, we used publicly available MVP Phecode-based genome-wide summary statistics to define sleep-related traits (Table S1).

##### All of Us

The All of Us Research Program is a longitudinal cohort study aimed at improving precision medicine by collecting genetic, lifestyle, and health data from a diverse U.S. population.^69^ In this study, we used publicly available summary statistics from sleep disorders as defined by the All by All project.^70^

##### Estonian Biobank

The Estonian Biobank is a population-based biobank with 212,955 participants (∼20% of the adult population).^71^ Health information on ICD-10 codes is obtained through regular linking with the National Health Insurance Fund and the National Electronic Health Record system using a unique national personal identification code. The majority of the electronic health records have been available since 2004, enabling integration of genetic data with longitudinal clinical information. Sleep-related phenotypes were defined using ICD-10 codes and analyzed independently (**Table S1**).

##### Meta-analysis

We performed fixed effects meta-analysis using METAL^72^ inverse variant weighted method. We performed clumping using plink v.2 using r^2^ cut-off of 0.2 against the UK Biobank European reference panel to identify independent association signals.

##### Fine mapping association signals

Fine-mapping was performed on all autosomal predefined HG38 genome-wide LD Blocks (https://github.com/jmacdon/LDblocks_GRCh38/blob/master/data/deCODE_EUR_LD_blocks.bed) with at least one or more variants that had a genome-wide significant P-value (P < 5 × 10^−8^) and passed a MAF filter of 0.01 using Sum of Single Effects (SuSiE) Regression on Summary Statistics.^73,74^ The R-package "susieR" was used to run the analysis, with the function susie_rss(), utilizing default parameters with R v.4.0.0. 10,000 randomly selected, unrelated participants of the MGB Biobank of European ancestry were used as the LD reference panel for the analysis. Previously identified insomnia risk loci were sourced from Watanabe et al.^75^, RLS from Winkelman et al.^76^ and Schormair et al.^38^, Narcolepsy from Han et al.^77^ and Ollila et al.^78^. Loci that did not overlap with any previously reported loci were considered novel findings.

##### Sleep related traits and medications genetic correlations and cohort heritability

Genetic correlations and observed heritability estimates were calculated using LD Score Regression (LDSC) version 1.0.1. Summary statistics were edited into the proper format using the munge_sumstats.py tool, and the variants were merged with the HapMap3 SNPs list to only include those within the LD reference panel. We excluded phenotypes with small sample sizes and retained only those with an observed SNP-based heritability greater than 0.03 and a standard error narrow enough to keep the 95% confidence interval between 0 and 1. In addition, we computed polygenic risk scores between sleep and neuropsychiatric traits using PRScs.

##### FUMA and MAGMA gene set and cell type enrichment analyses

We used FUMA and MAGMA gene set and cell type enrichment analyses to examine enrichment of pathways and tissue specific expression across the tested traits.^75,79^

##### Disease Endpoint PheWAS and expression-qtl and protein-qtl annotation of individual variants

We performed pheWAS by testing individual variants for association across a broad range of disease endpoints in the FinnGen, and publicly available datasets. For variants previously reported as eQTLs or pQTLs, we assessed their trait associations in these cohorts.

##### Bayesian clustering of effects based on linear relationships

We compared effect size estimates between sleep and comorbid traits by selecting genome-wide significant variants for trait pair. The line models R-package was utilized for comparing linear relationships (https://github.com/mjpirinen/linemodels). This line model method performs probabilistic clustering of variables based on their observed effect sizes on two outcomes.^80,81^

##### Mendelian randomization analysis

To investigate potential causal relationships between traits, we performed Mendelian randomization (MR) using the TwoSampleMR package in R.^82,83^ Genetic instruments were derived from genome-wide significant variants (p < 5 × 10LL), clumped for independence at r² < 0.01 within a 10 Mb window, using the European reference panel from 1000 Genomes Project. We applied the inverse-variance weighted (IVW) method as the primary estimator, which provides an efficient test under the assumption that all instruments are valid. To account for possible violations of MR assumptions due to horizontal pleiotropy, we complemented the IVW approach with MR-Egger regression.^84^ The MR-Egger intercept was used to test for directional pleiotropy, with a significant non-zero intercept indicating potential bias. We further assessed robustness by performing weighted median and mode-based estimators^84^, which provide consistent causal estimates even when up to 50% of instruments are invalid. All MR analyses were harmonized to align effect alleles across exposure and outcome datasets, and summary statistics were harmonized and formatted according to the MRC IEU OpenGWAS standards.^85,86^ We used publicly available data from FinnGen laboratory value GWAS for ferritin levels for exposure variables.

##### Ethics Statements

###### FinnGen

Patients and control subjects in FinnGen provided informed consent for biobank research, based on the Finnish Biobank Act. Alternatively, separate research cohorts, collected prior the Finnish Biobank Act came into effect (in September 2013) and start of FinnGen (August 2017), were collected based on study-specific consents and later transferred to the Finnish biobanks after approval by Fimea (Finnish Medicines Agency), the National Supervisory Authority for Welfare and Health. Recruitment protocols followed the biobank protocols approved by Fimea. The Coordinating Ethics Committee of the Hospital District of Helsinki and Uusimaa (HUS) statement number for the FinnGen study is Nr HUS/990/2017.

The FinnGen study is approved by Finnish Institute for Health and Welfare (permit numbers: THL/2031/6.02.00/2017, THL/1101/5.05.00/2017, THL/341/6.02.00/2018, THL/2222/6.02.00/2018, THL/283/6.02.00/2019, THL/1721/5.05.00/2019 and THL/1524/5.05.00/2020), Digital and Population Data Service Agency (permit numbers: VRK43431/2017–3, VRK/6909/2018–3, VRK/4415/2019–3), the Social Insurance Institution (permit numbers: KELA 58/522/2017, KELA 131/522/2018,KELA 70/522/2019, KELA 98/522/2019, KELA 134/522/2019, KELA 138/522/2019, KELA 2/522/2020,KELA 16/522/2020), Findata (permit numbers: THL/2364/14.02/2020, THL/4055/14.06.00/2020, THL/3433/14.06.00/2020, THL/4432/14.06/2020, THL/5189/14.06/2020, THL/5894/14.06.00/2020, THL/6619/14.06.00/2020, THL/209/14.06.00/2021, THL/688/14.06.00/2021, THL/1284/14.06.00/2021, THL/1965/14.06.00/2021, THL/5546/14.02.00/2020) and Statistics Finland (permit numbers: TK-53-1041-17 and TK/143/07.03.00/2020 (earlier TK-53-90-20)).

The Biobank Access Decisions for FinnGen samples and data utilized in FinnGen Data Freeze7 include: THL Biobank BB2017_55, BB2017_111, BB2018_19, BB_2018_34, BB_2018_67, BB2018_71, BB2019_7, BB2019_8, BB2019_26, BB2020_1, Finnish Red Cross Blood Service Biobank 7.12.2017, Helsinki Biobank HUS/359/2017, Auria Biobank AB17-5154 and amendment number1 (August 17 2020), Biobank Borealis of Northern Finland_2017_1013, Biobank of Eastern Finland 1186/2018 and amendment 22/2020, Finnish Clinical Biobank Tampere MH0004 and amendments (21.02.2020, 06.10.2020), Central Finland Biobank 1–2017, and Terveystalo Biobank STB 2018001.2.

##### UK Biobank

The North West Multi-centre Research Ethics Committee (MREC) has granted the Research Tissue Bank (RTB) approval for the UKB that covers the collection and distribution of data and samples (http://www.ukbiobank.ac.uk/ethics/). Our work was performed under the UKB application number 22627 and 9072. All participants included in the conducted analyses have given a written consent to participate.

##### MGB Biobank

The MGB Biobank has obtained a Certificate of Confidentiality. In addition, The MGB Biobank works in close collaboration with the Partners Human Research Committee (PHRC) (the Institutional Review Board). This collaboration has ensured that the Biobank’s actions and procedures meet the ethical standards for research with human subjects. Biobank patients are recruited from inpatient stays, emergency department settings, outpatient visits, and electronically through a secure online portal for patients. Recruitment and consent materials are fully translated in Spanish to promote patient inclusion. The systematic enrollment of patients across the MGB network and the active inclusion of patients from diverse backgrounds contribute to a Biobank reflective of the overall demographic of the population receiving care within the MGB network. Recruitment for the Biobank launched in 2009 and is ongoing through both in-person recruitment at participating clinics and electronically through the patient portal. The recruitment strategy has been described previously.^61^ All recruited patients provided written consent upon enrollment, and are offered an option to refuse consent.

##### All of Us

The All of Us Research Program is a nationwide longitudinal cohort study approved by centralized institutional review board (IRB) oversight coordinated by the U.S. National Institutes of Health. All participants provided written informed consent for the collection and use of genetic, lifestyle, and health data in biomedical research from a diverse U.S. population.^69^ In this study, we extracted and analyzed publicly available, de-identified summary statistics from the All of Us data release (data freeze 2022) for sleep disorders across all ancestries (AFR, AMR, EAS, and EUR).

##### Million Veteran Program

The Million Veteran Project (MVP) is a national research initiative approved by the U.S Department of Veterans Affairs Central IRB. All participants provided written informed consent for the collection and use of their genetics, lifestyle, military experiences, and exposures affect health and wellness, including linkage with electronic health records.^67,68^ In this study, we extracted and analyzed publicly available, de-identified summary statistics from MVP (data freeze 4) for sleep disorders for all ancestries (AFR, AMR, EAS, and EUR).

##### Estonian biobank

The activities of the EstBB are regulated by the Human Genes Research Act, which was adopted in 2000 specifically for the operations of the EstBB. All participants provided broad informed consent for the use of their data and biological samples in health research. Individual level data analyses were conducted under ethical approval 1.1–12/624 from the Estonian Committee on Bioethics and Human Research (Estonian Ministry of Social Affairs), with data accessed under release application 6–7/GI/18324 from the Estonian Biobank.

## Data availability

Summary statistics will be made publicly available upon publication, both as supplementary data with the manuscript and through deposition in the Sleep Disorders Knowledge Portal (SDKP).

## Acknowledgements

This work was supported by the NIH grants R35 GM146839 (J.M.L), R01 HG012810 (J.M.L; H.M.O), R01 AI170850 (R.S.; H.M.O.) and R01 DK107859 (R.S). We acknowledge Research Council of Finland funding for H.M.O (350181). We gratefully acknowledge Biogen, the Estonian Biobank, FinnGen cohort, UK Biobank, the MGB Biobank, All of Us, and the Million Veteran Program, together with their investigators, staff, and affiliated institutions, for their essential contributions to data collection, and curation. We also thank colleagues who provided valuable feedback on the manuscript. Most importantly, we acknowledge the participants and their families, whose willingness to contribute personal health and genetic information underpins these biobanks and makes this research possible.

## Author contributions

R. S., J. M. L. and H. M. O. conceived the study and supervised the project. J. V., L. K., C. C. and M. M. performed the analyses. All authors contributed to the interpretation of the results. L. K and H.M.O drafted the manuscript with input from all authors. All authors reviewed and approved the final version of the manuscript.

## Competing interests

The authors declare no competing interests.

## Notes

### Competing Interest Statement

The authors have declared no competing interest.

### Author Declarations

Data used is from the UK biobank, MGB biobank, Million Veterans Program, FinnGen cohort, All of Us Biobank, and Estonian Biobank

