## Supplement for "From Normal Variation in Sleep to Clinical Sleep Disorders: Genetic Insights from Over One Million Individuals"

**Table of contents.**

1. **List of Supplementary tables and table legends**

**Table S1.** Cohort description

**Table S2.** Demographics

**Table S3.** Finemapped meta-analysis lead variants

**Table S4.** Heritability estimates

**Table S5.** Replication in the Estonian Biobank

**Table S6** Transcription factor targets in Sleep apnea

**Table S7** Between trait associations**.**

**Table S8.** Variant GWAS pheWAS annotation from open targets, FinnGen, GWAS Catalog,

**Table S9.** List of medications used in the study

**Table S10**. Earlier reported genome-wide associations with clinical sleep traits

**Table S11.** Genetic correlation and PRS analysis

**Table S12.** Mendelian Randomization

**Table S13.** FinnGer author banner

2. List of supplementary Figures

**Figure S1** Manhattan and QQ plots of all studied traits

**Figure S2** Trait heritability estimates

**Figure S3** Functional enrichment in clinical insomnia

**Figure S4** Functional enrichment in RLS

**Figure S5** Functional enrichment in sleep apnea

**Figure S6** Genetic correlation between sleep
and selected neuropsychiatric diseases

**Figure S7** Mendelian randomization

**Figure S1a** ICD_all_sleep

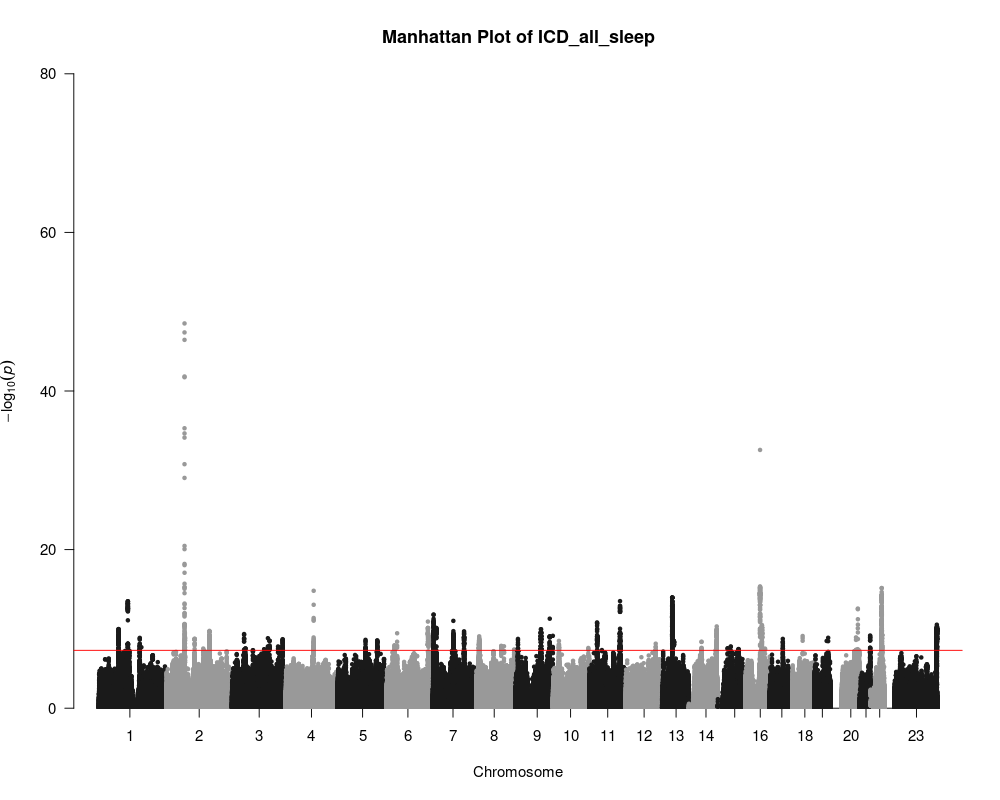

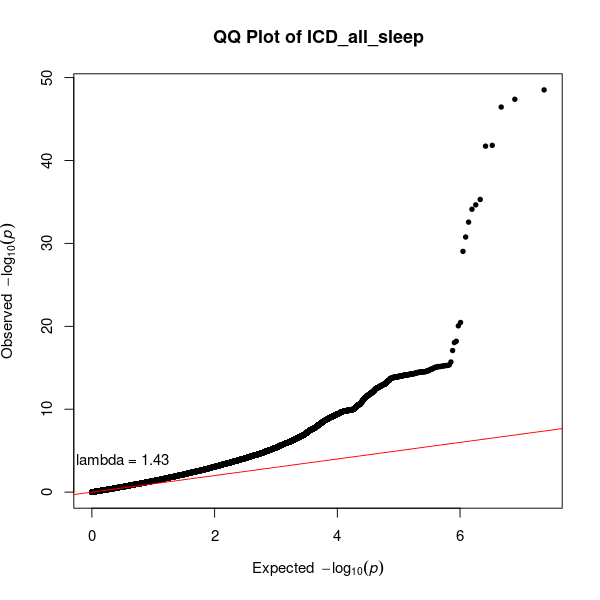

**Figure S1b** Clinical insomnia

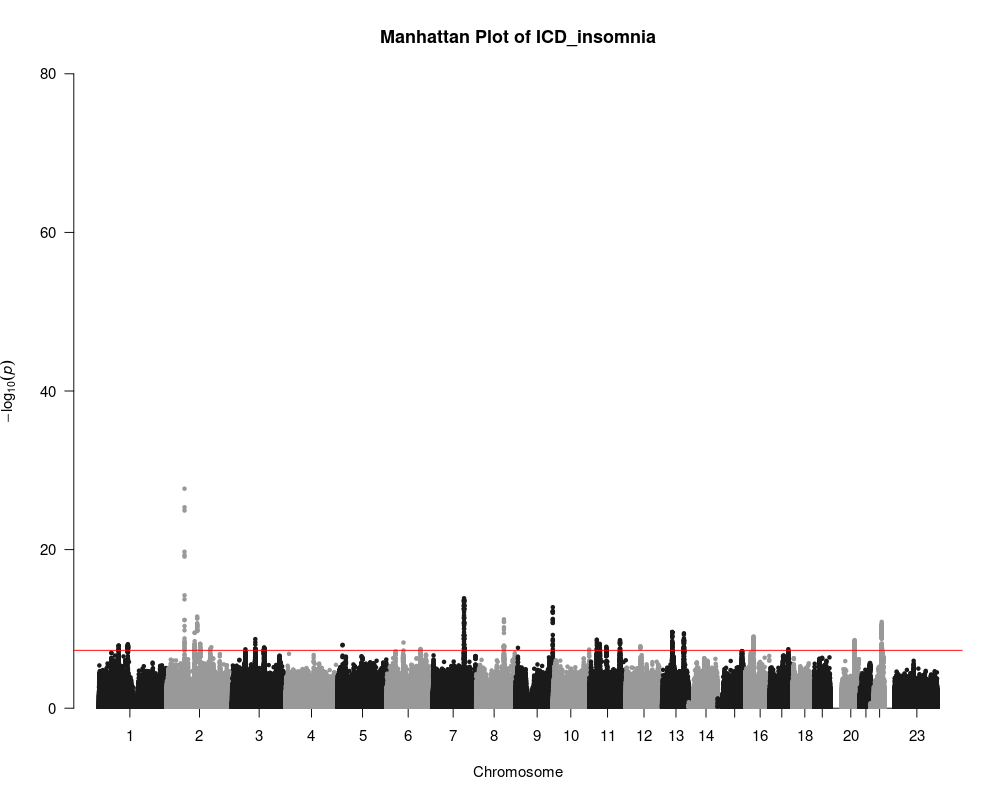

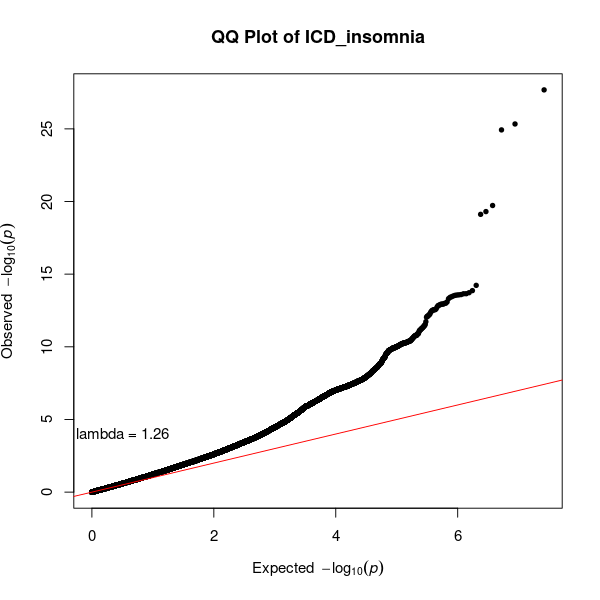

**Figure S1c** Hypersomnia

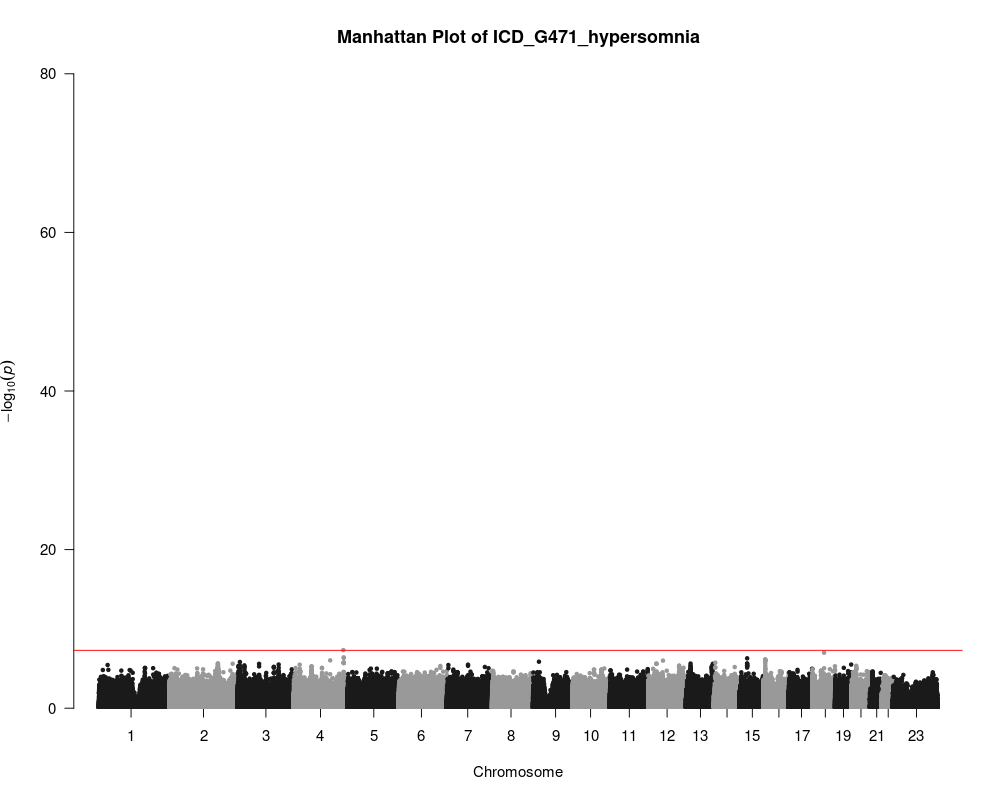

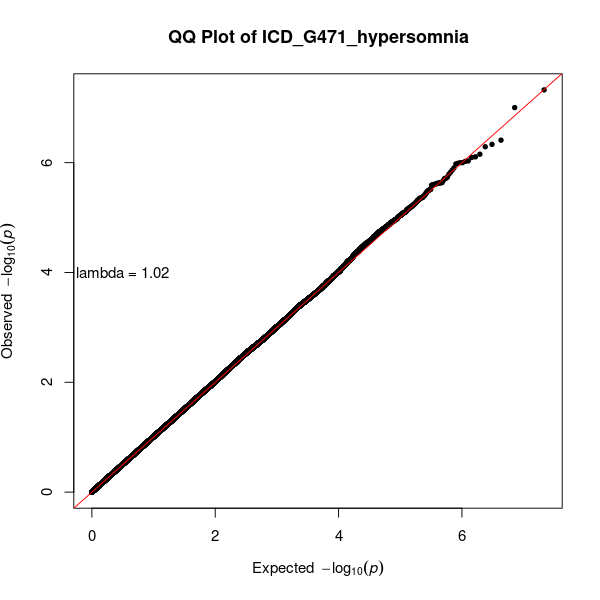

**Figure 1d.** ICD_apnea

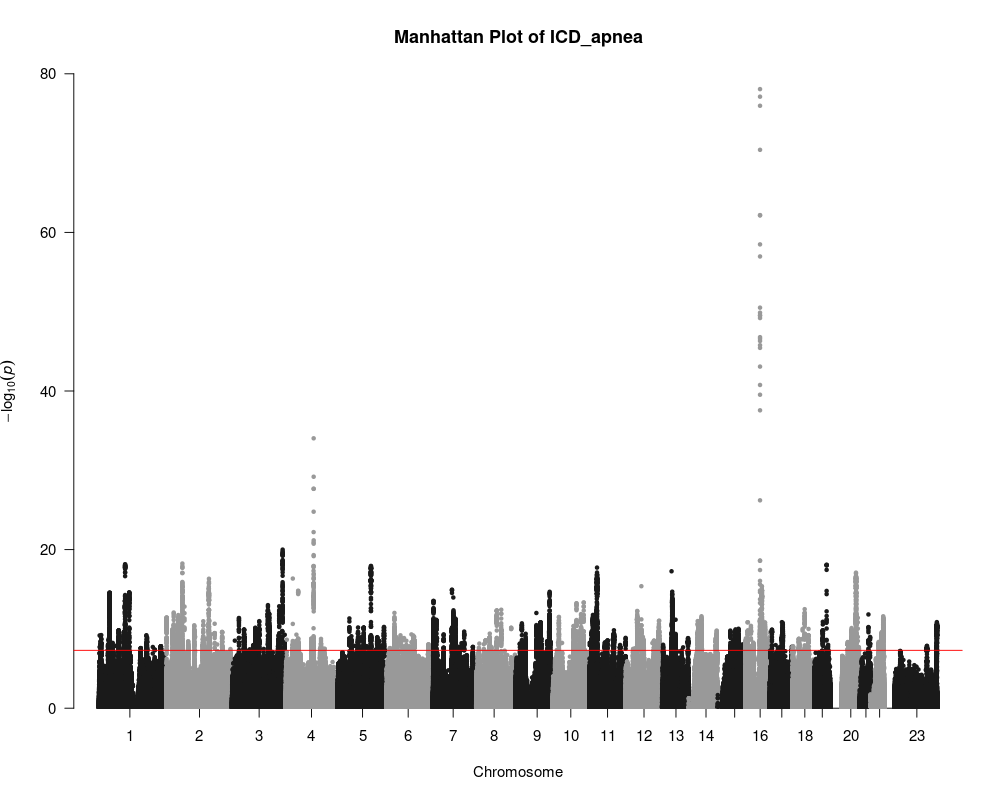

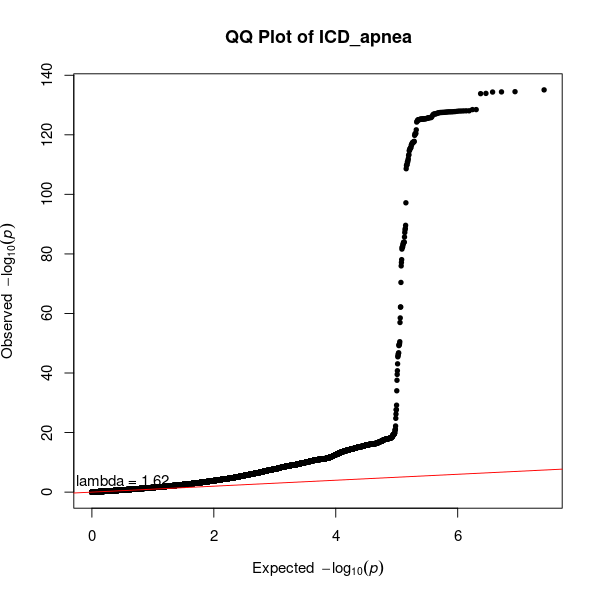

**Figure 1e.** ICD_restlessleg

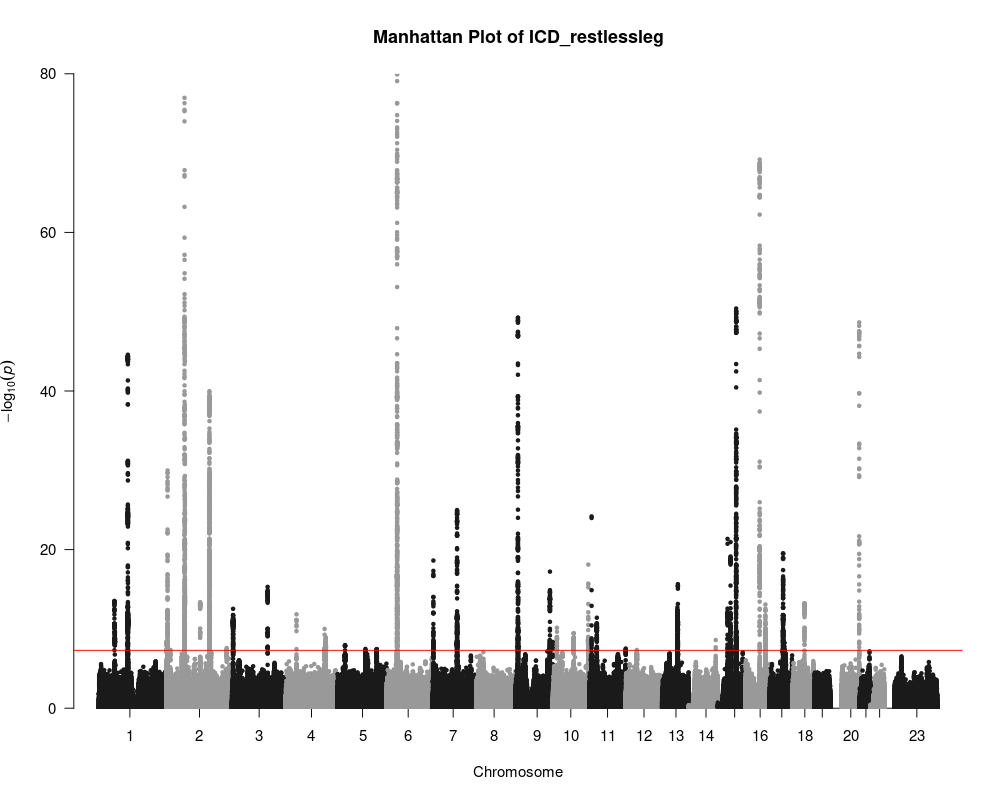

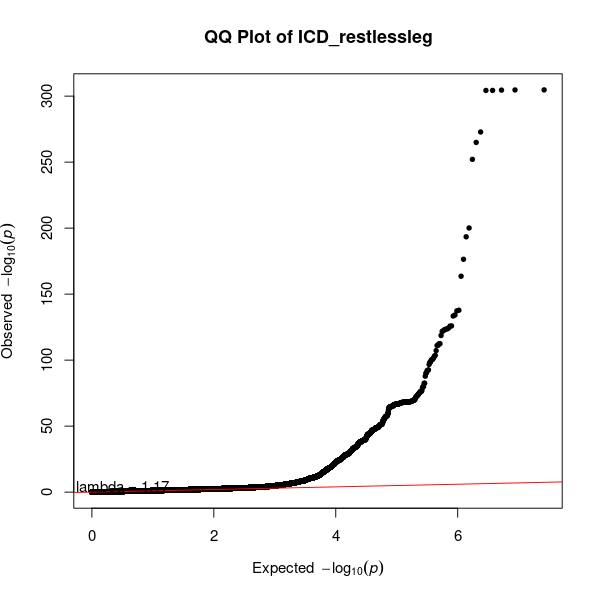

**Figure 1f**. ICD_G933_fatigue

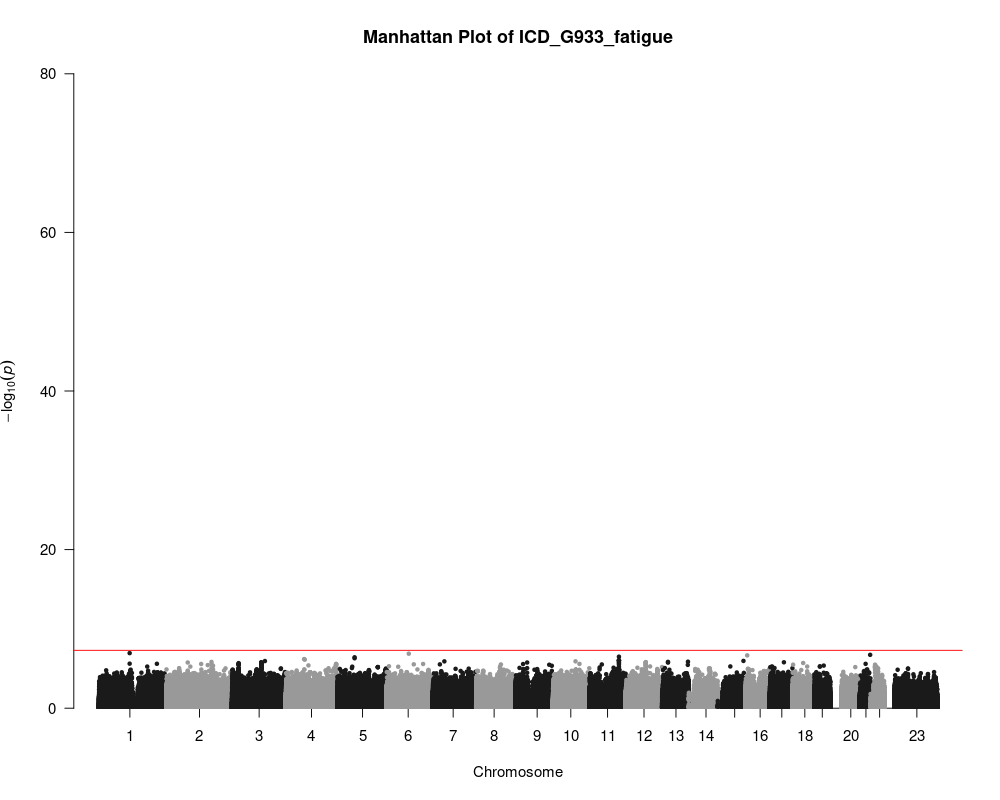

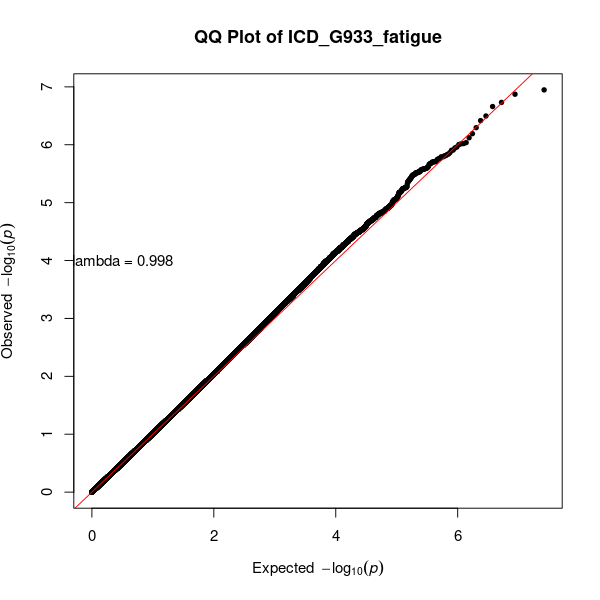

**Figure 1g.** ICD_narcolepsy

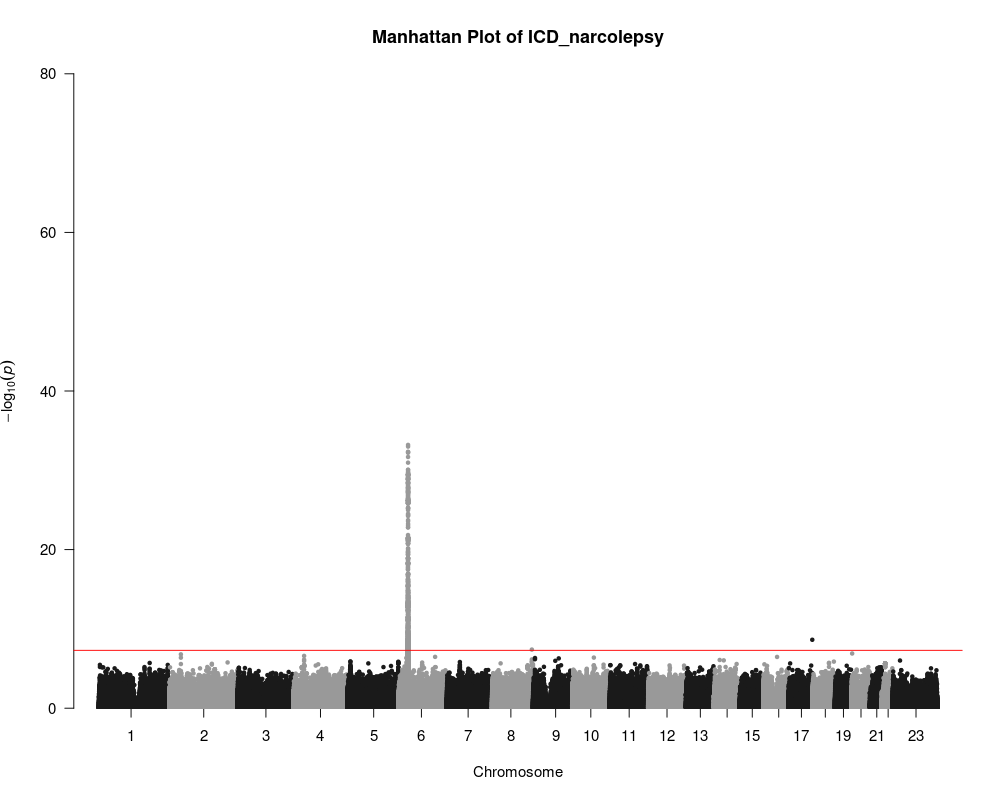

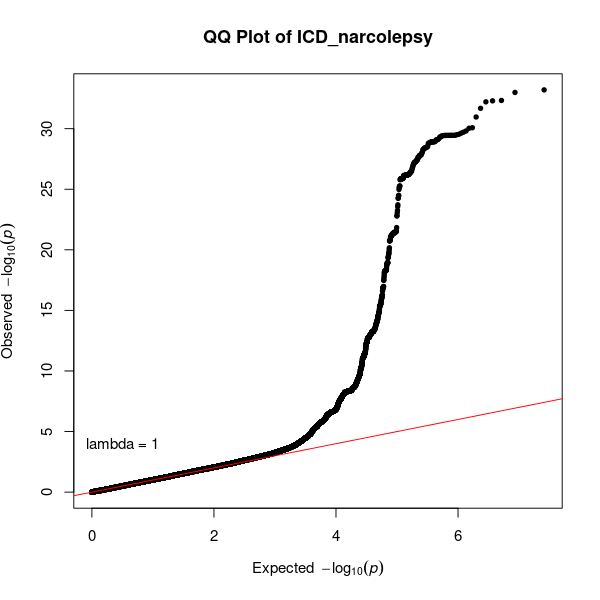

**Figure 1h**. MEDS_all

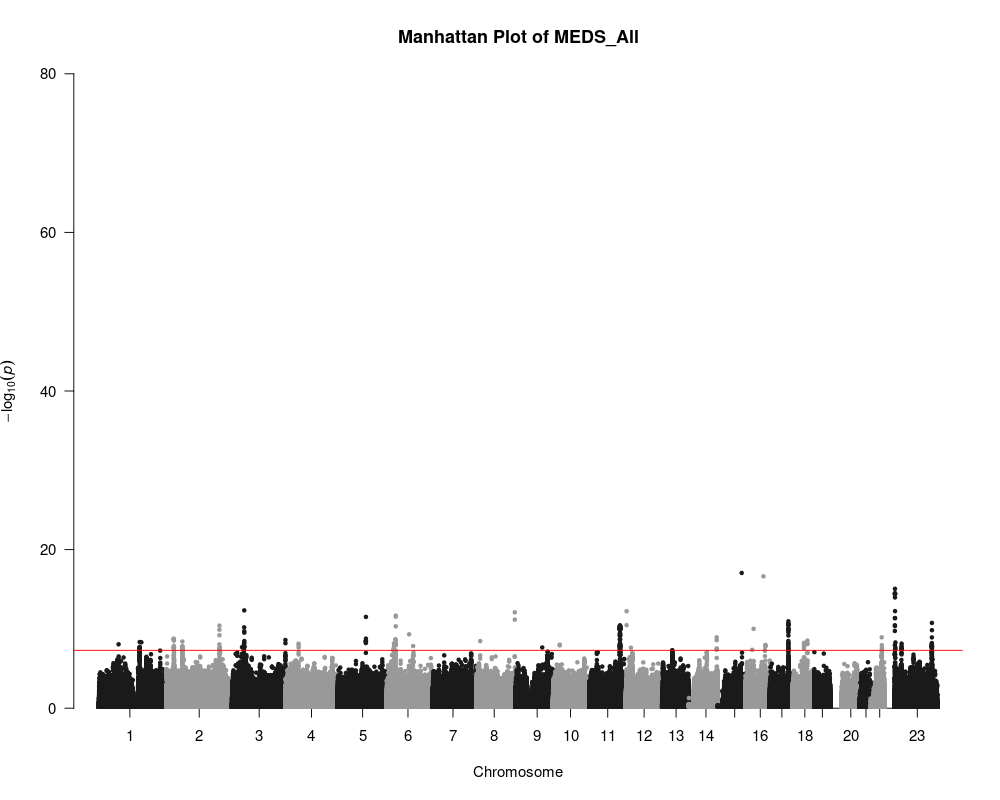

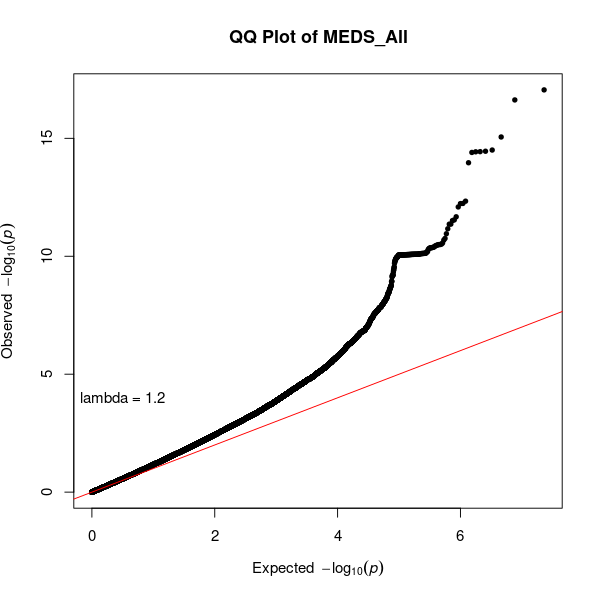

**Figure 1i.** MEDS_insomnia_circadian_only

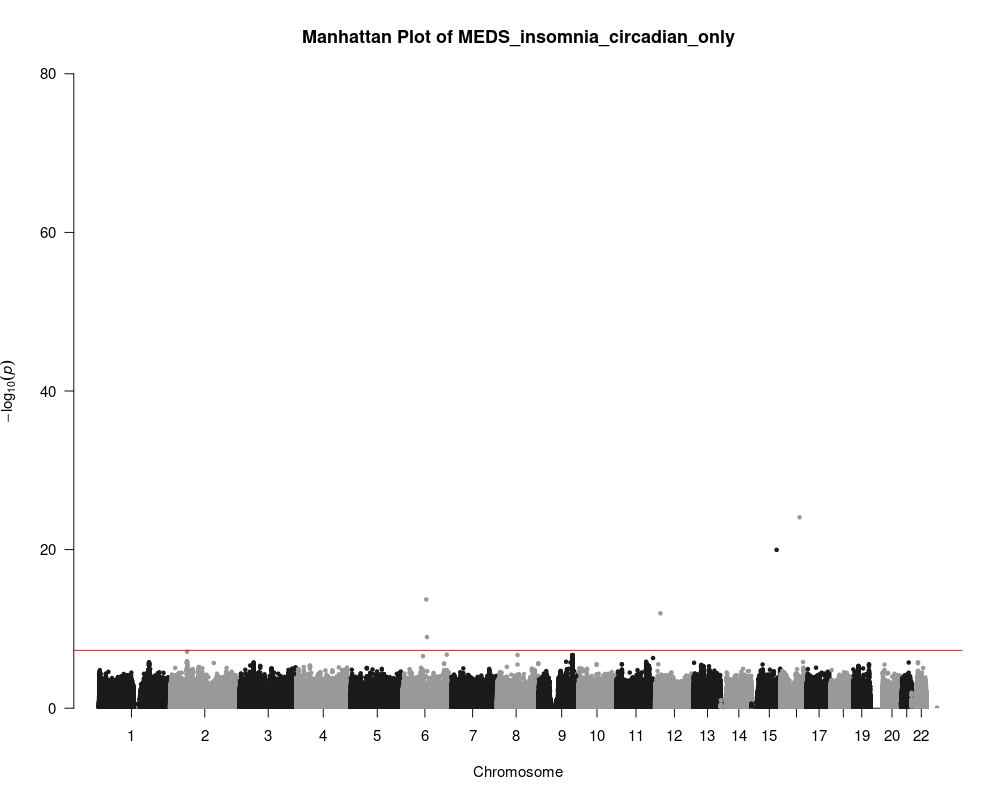

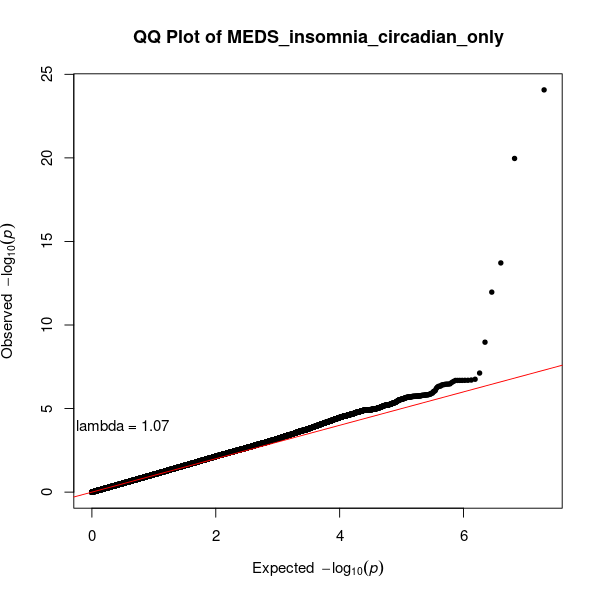

**Figure 1j.** MEDS_anxiety_only

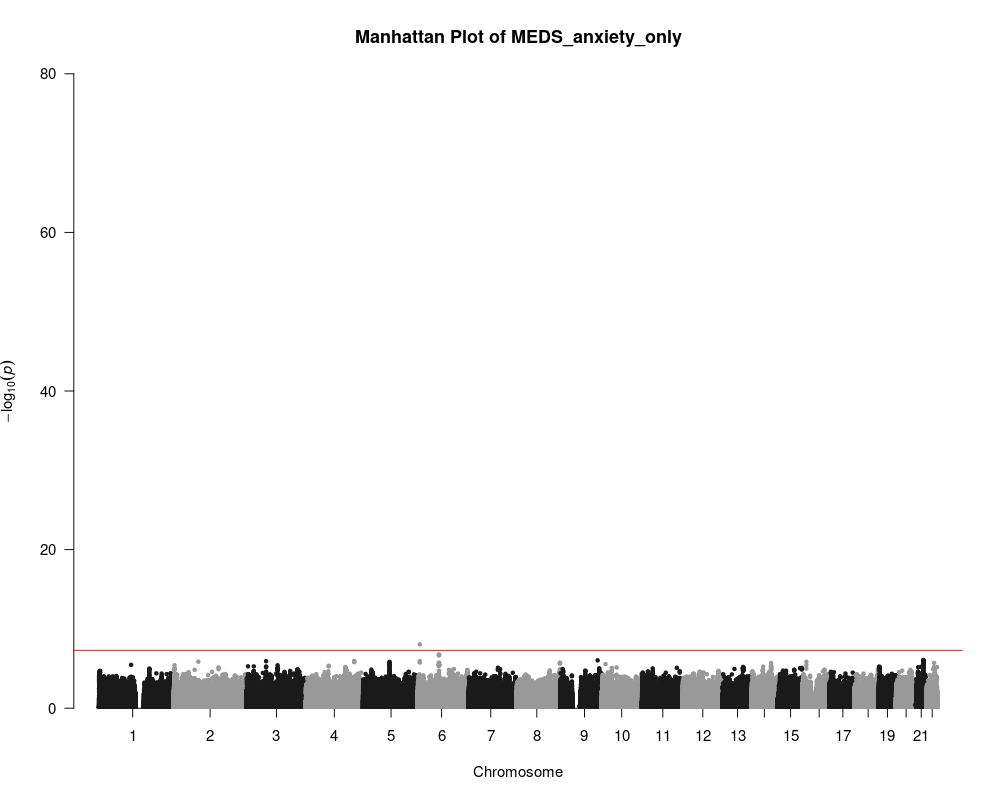

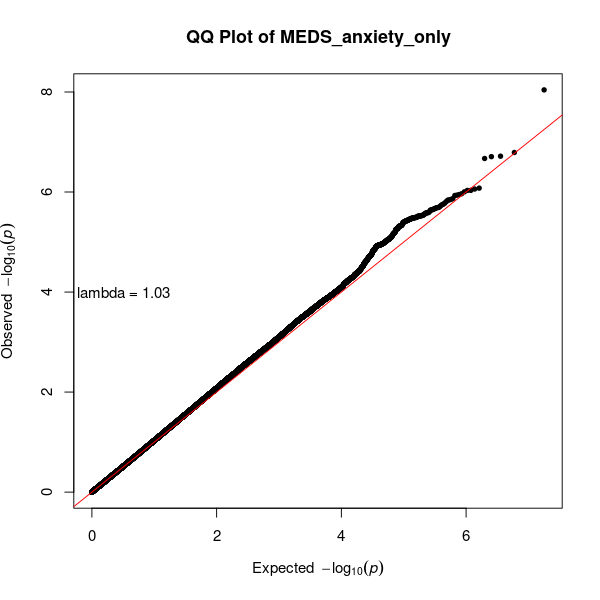

**Figure 1k.** MEDS_insomnia_only

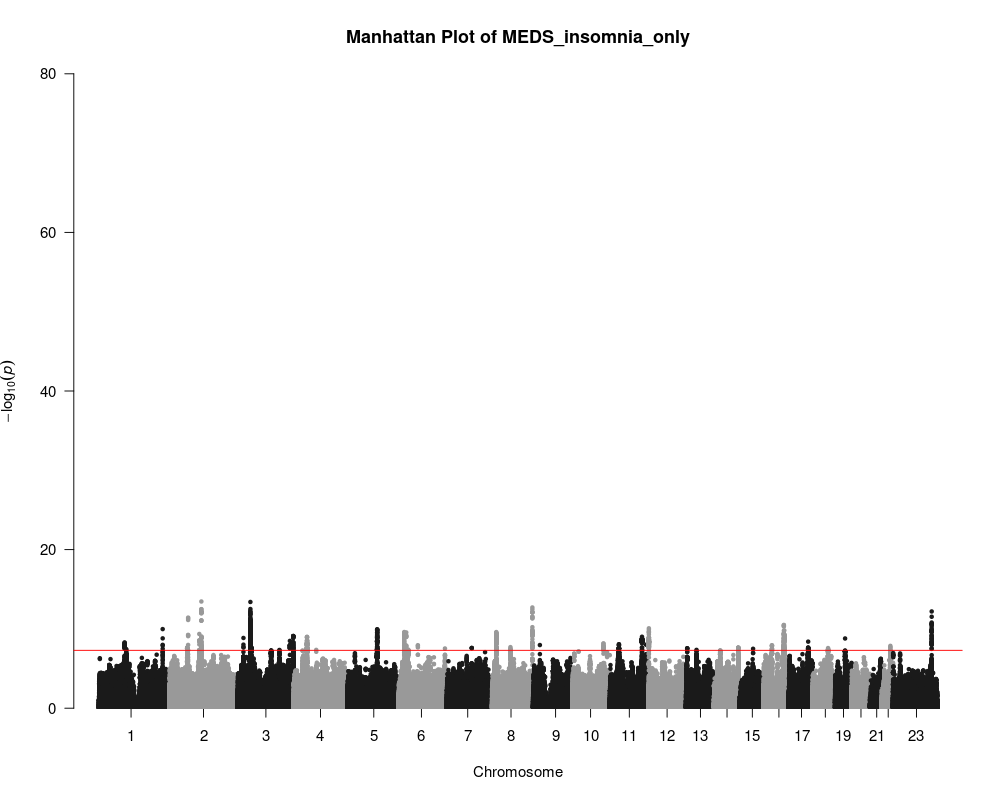

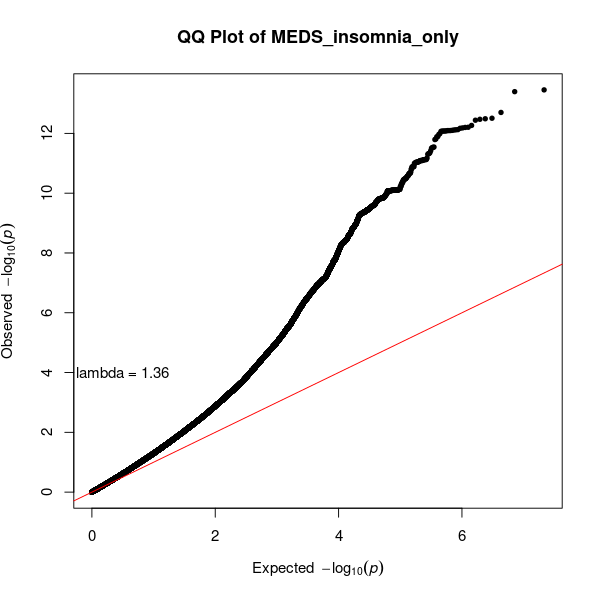

**Figure 1l.** MEDS_chlordiazepoxide

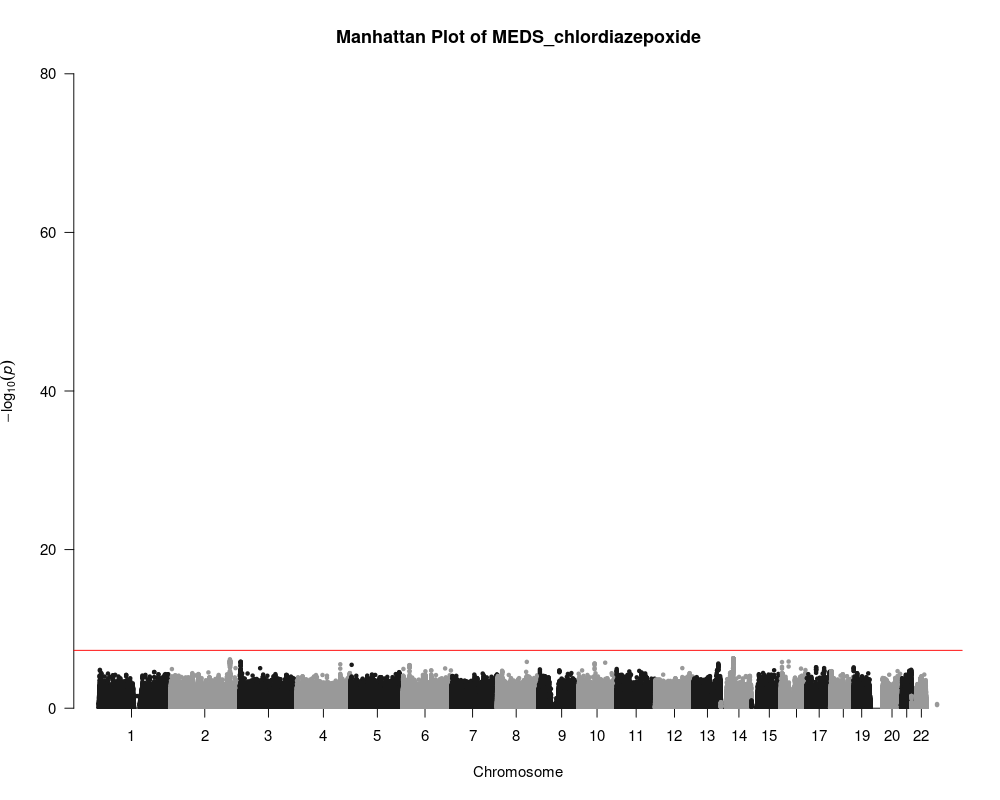

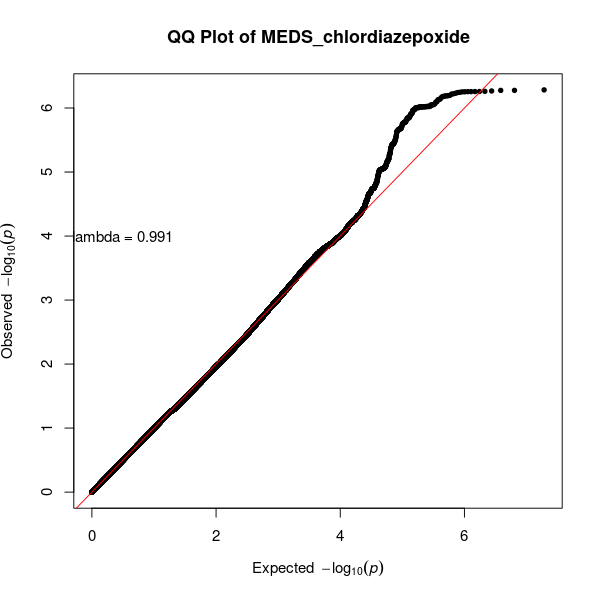

**Figure 1m**. MEDS_diazepam

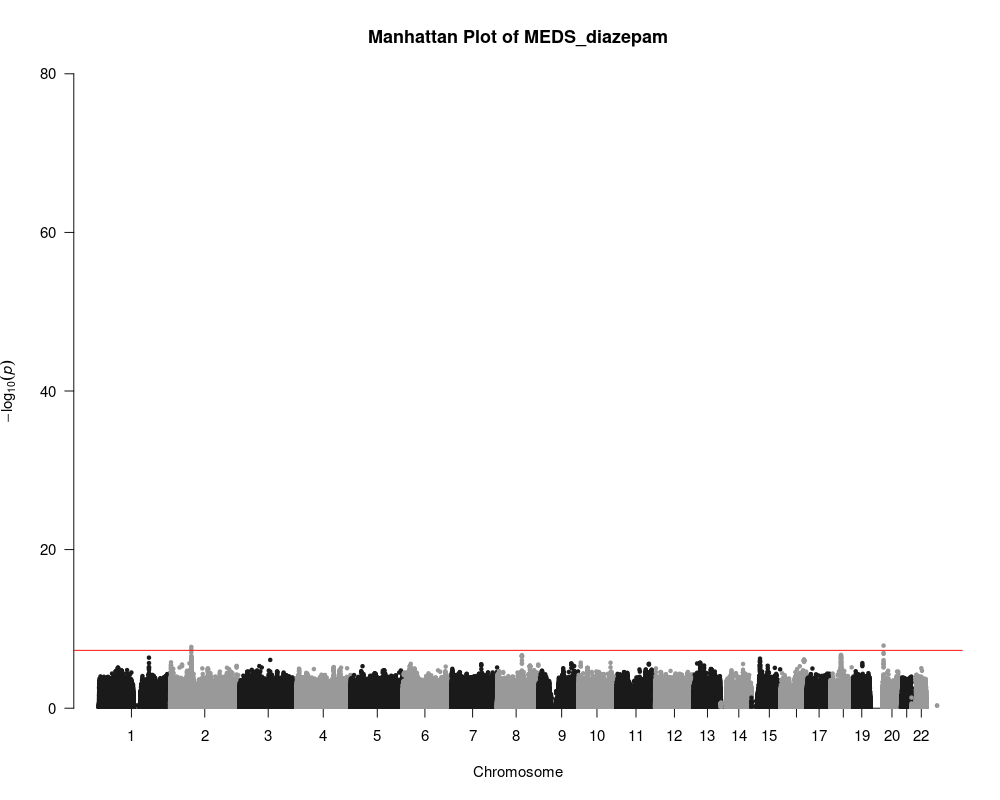

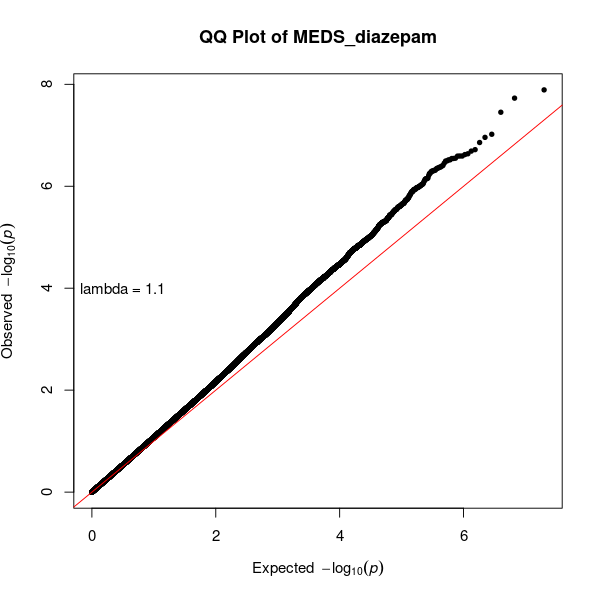

**Figure 1n**. MEDS_hydroxyzine

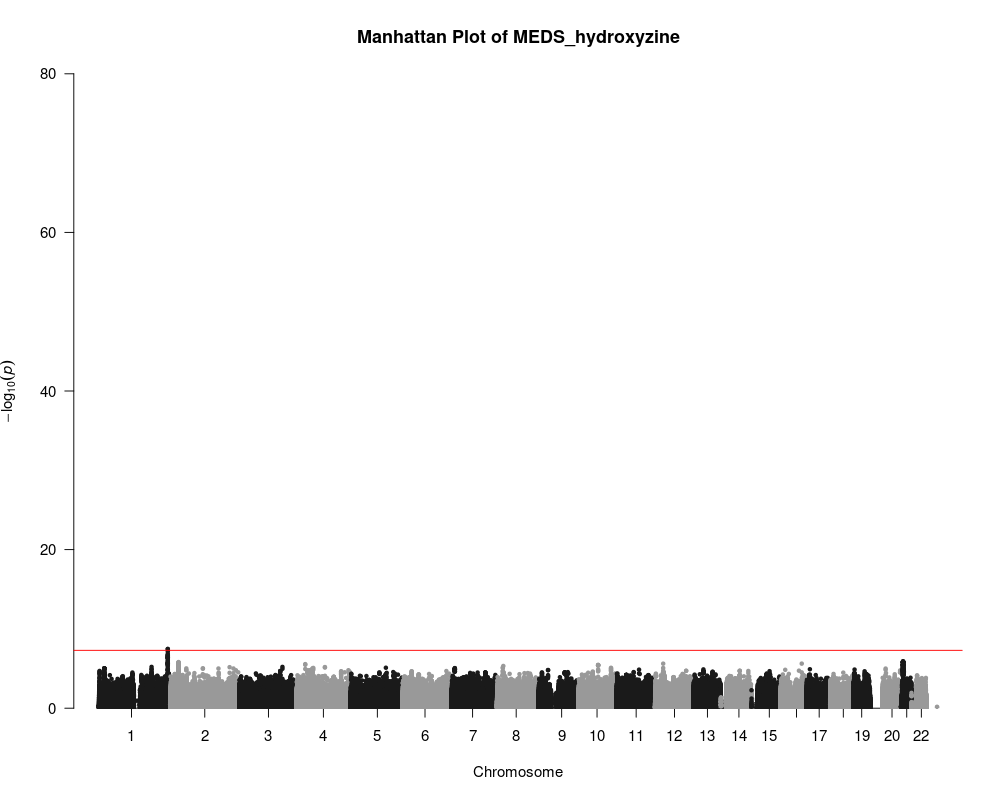

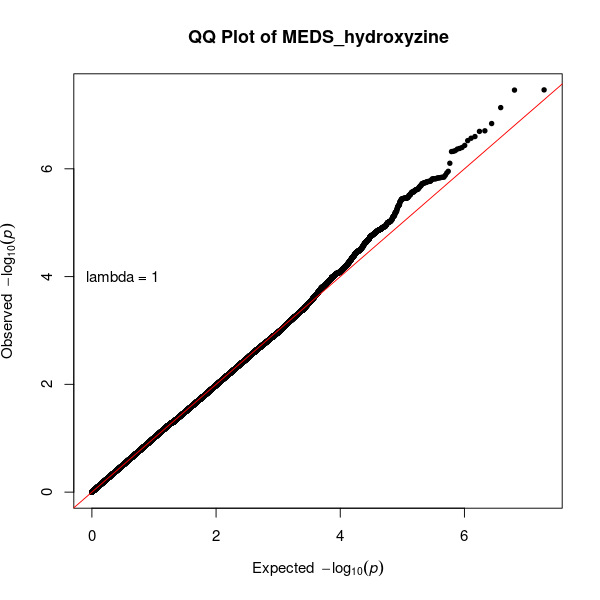

**Figure 1o.** MEDS_lorazepam

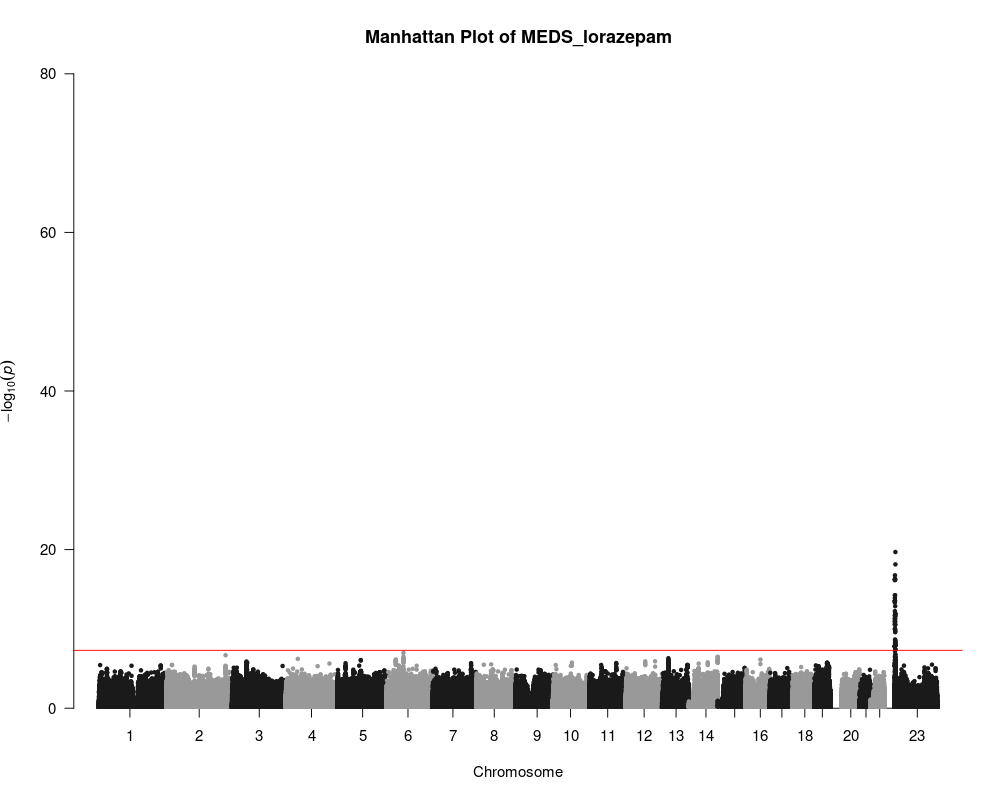

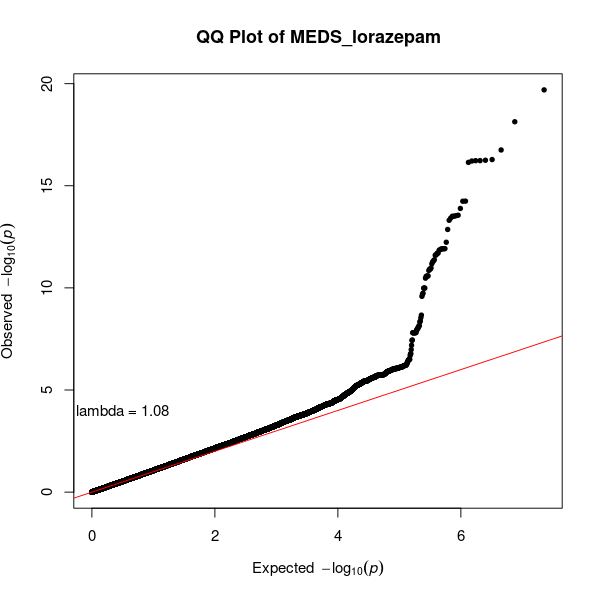

**Figure 1p.** MEDS_oxazepam

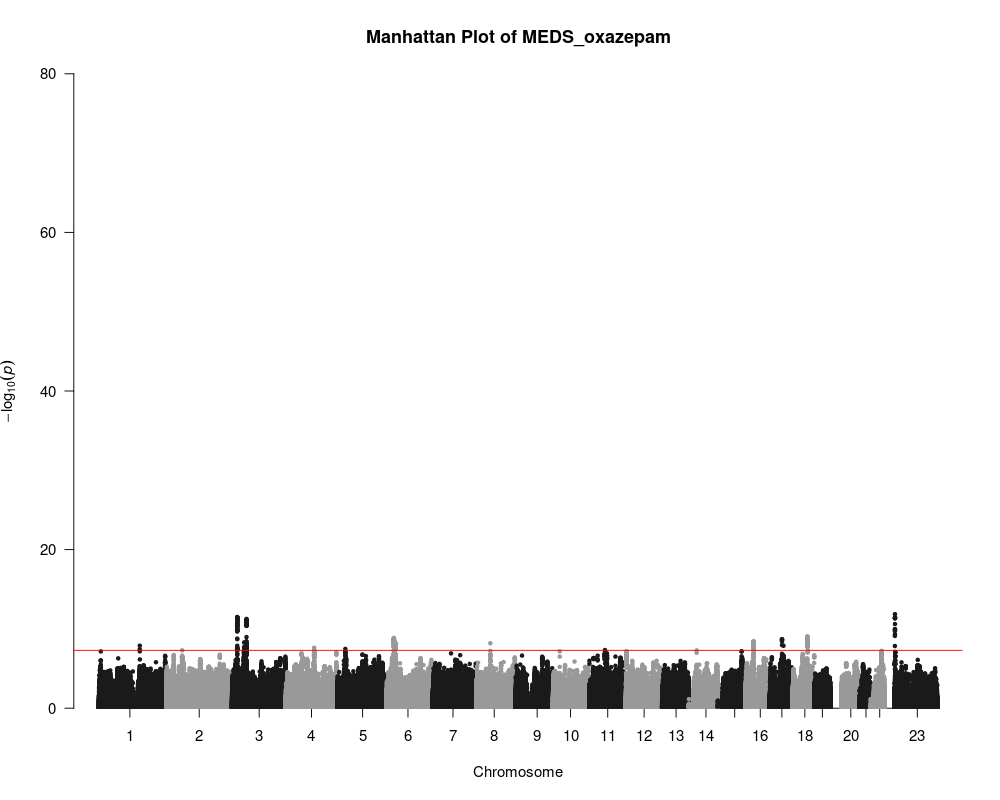

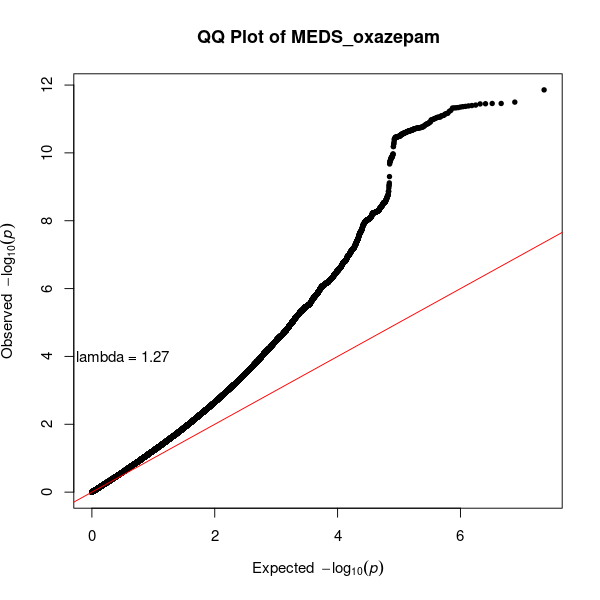

**Figure 1q**. MEDS_temazepam

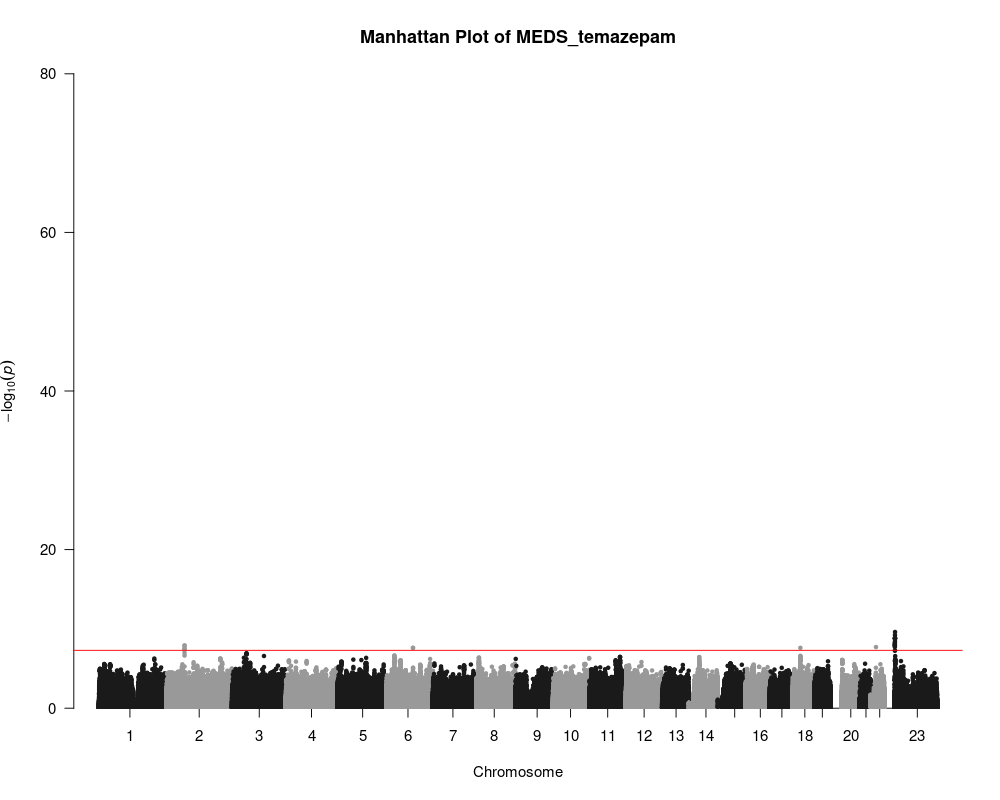

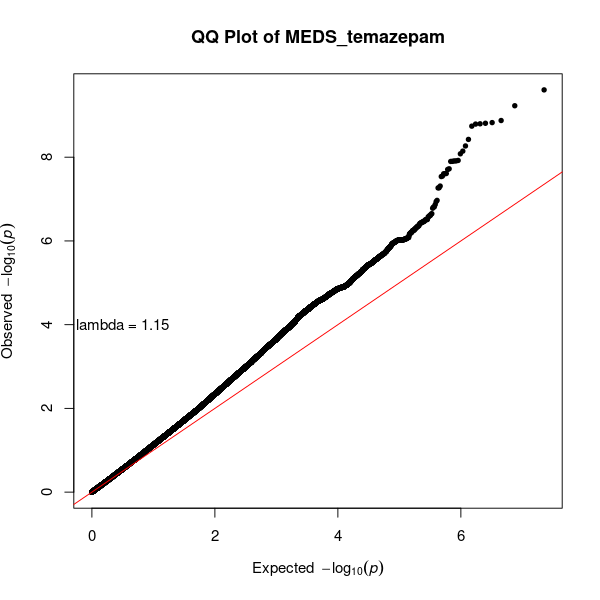

**Figure 1r.** MEDS_zaleplon

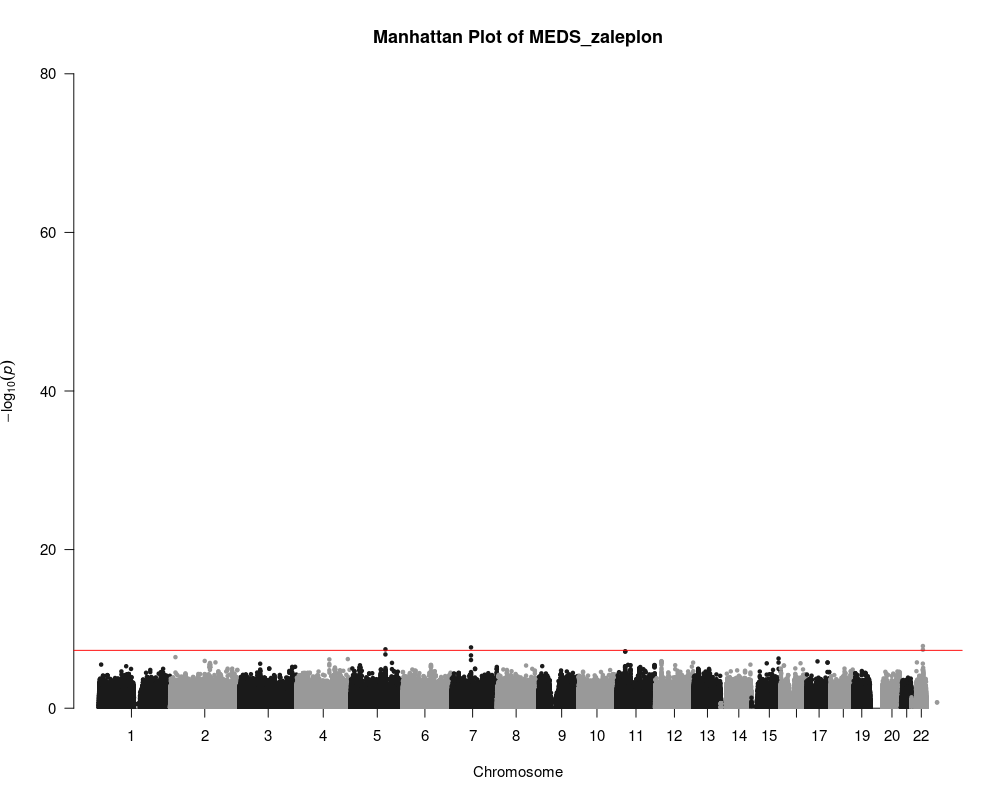

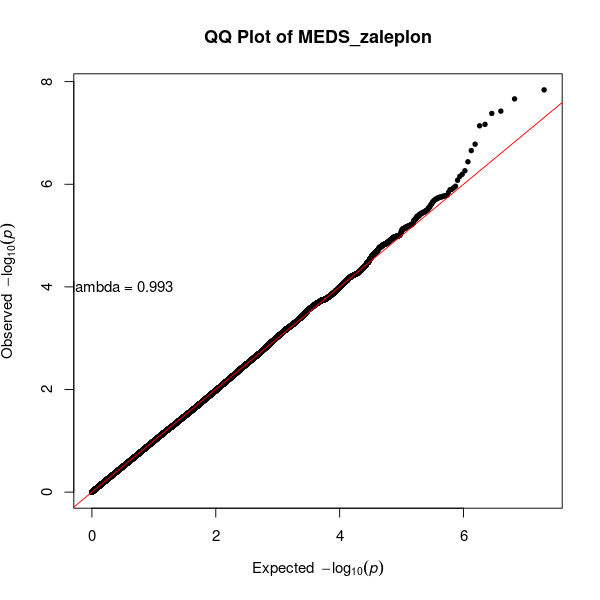

**Figure 1s**. MEDS_zolpidem

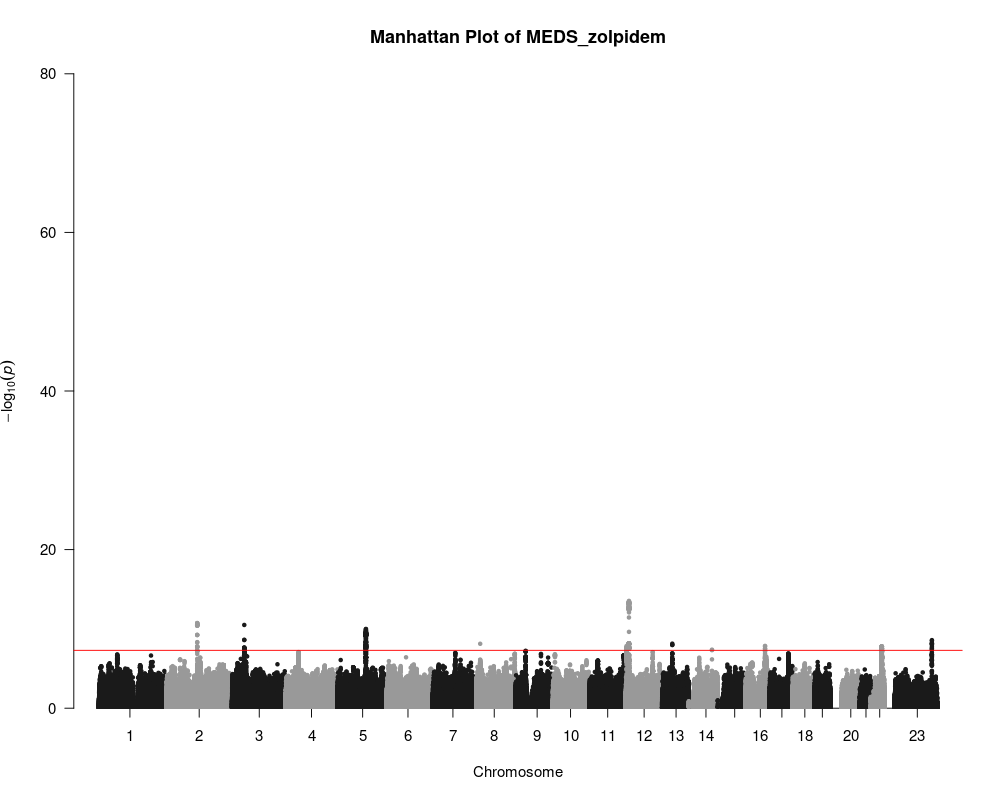

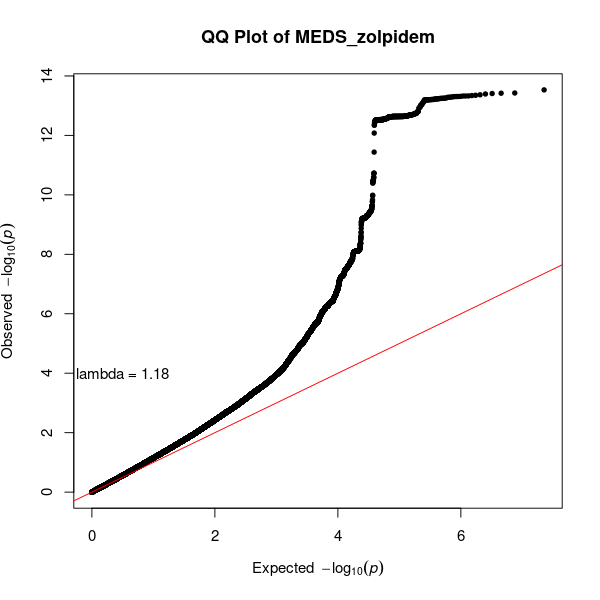

**Figure 1t.** MEDS_zopiclone

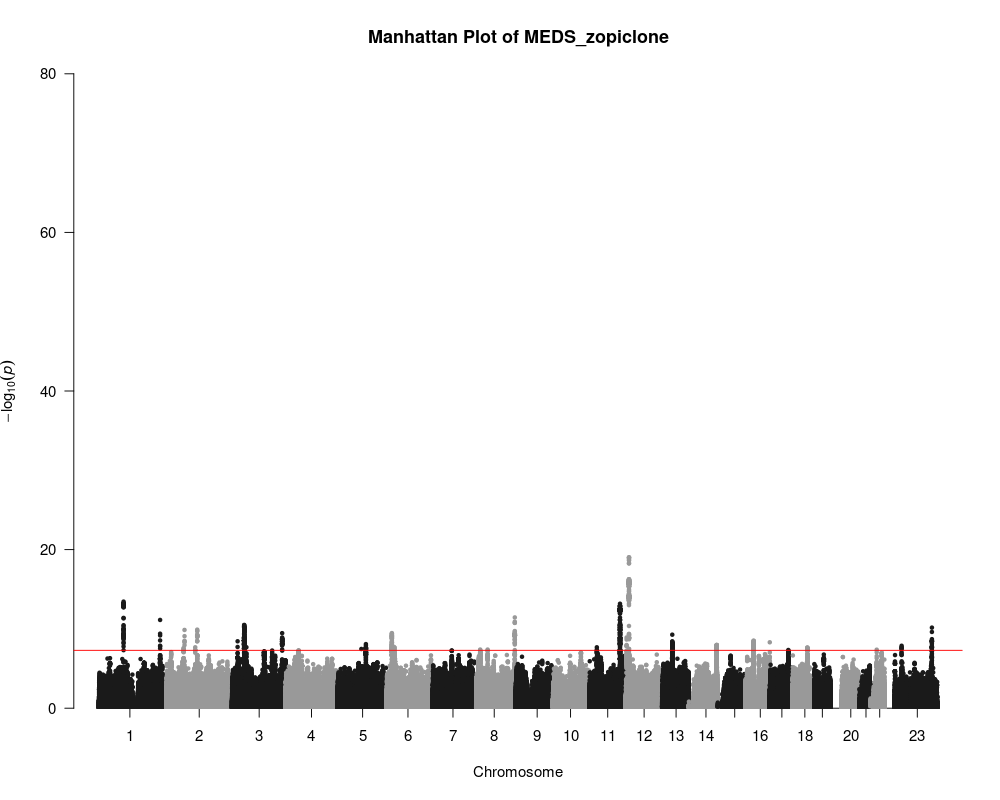

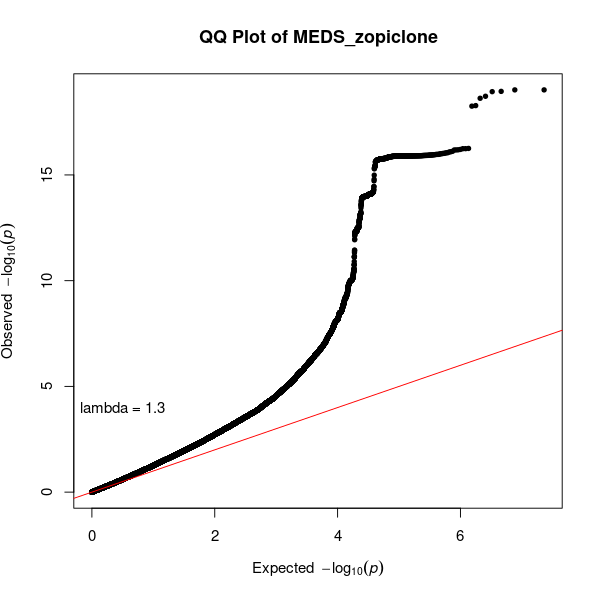

**Figure 1u**. MEDS_clobazam

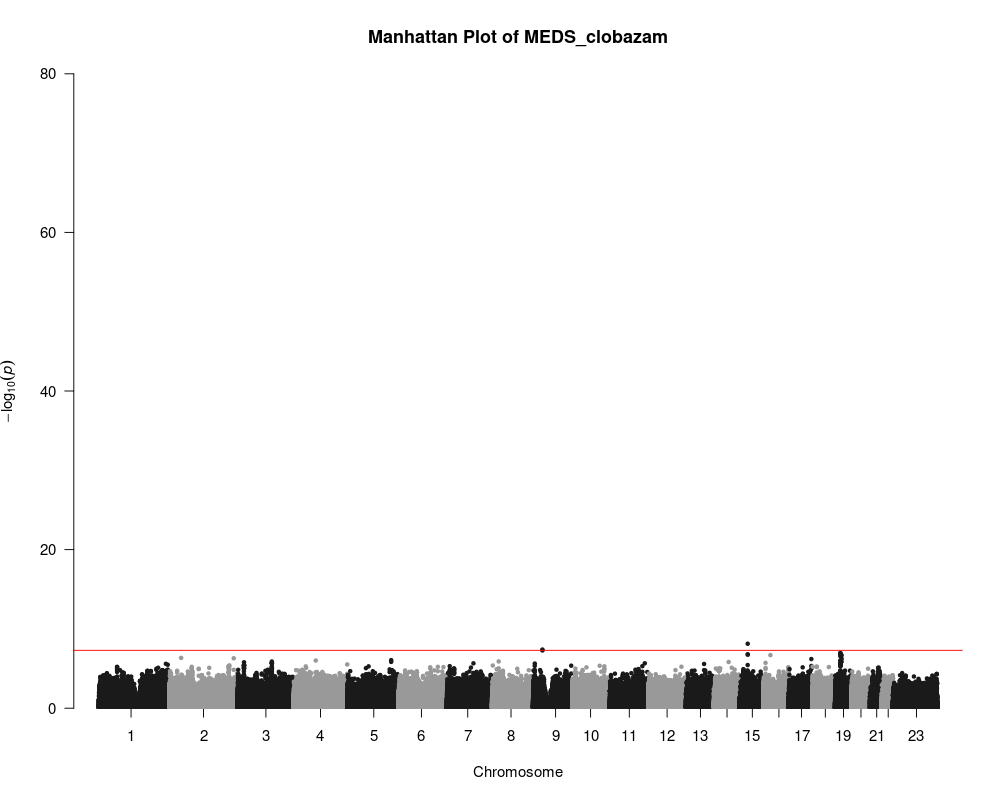

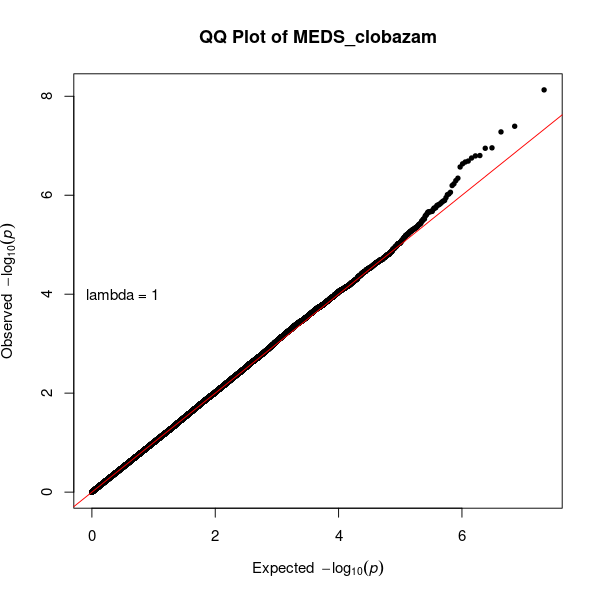

**Figure 1v.** MEDS_triazolum

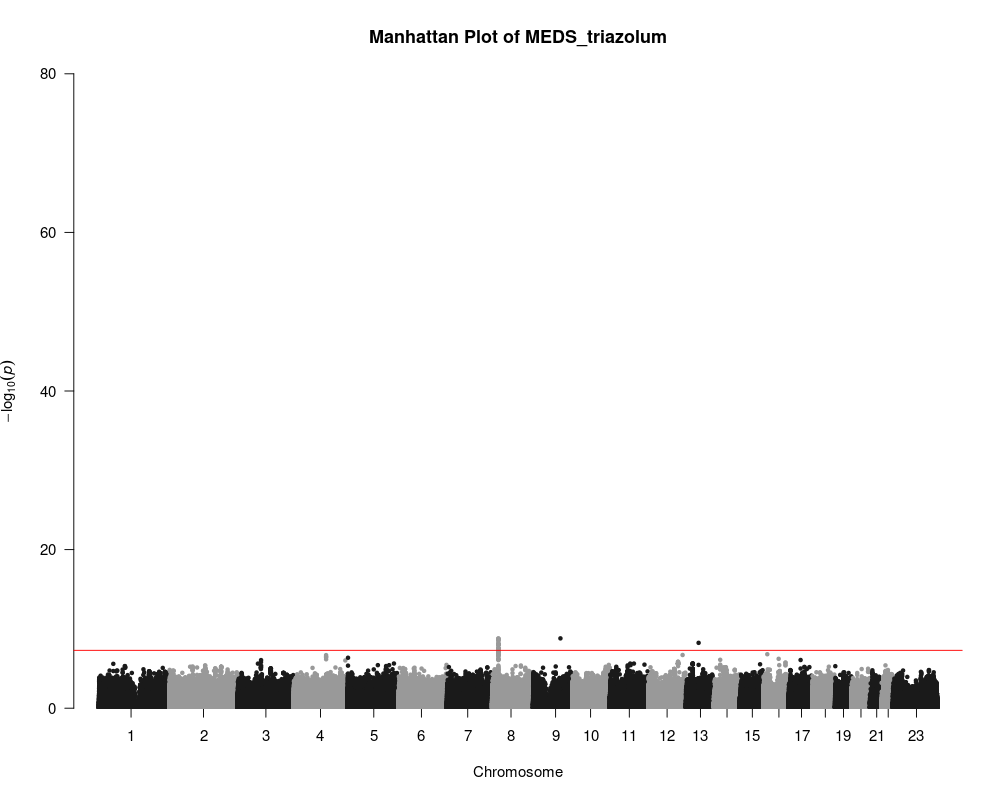

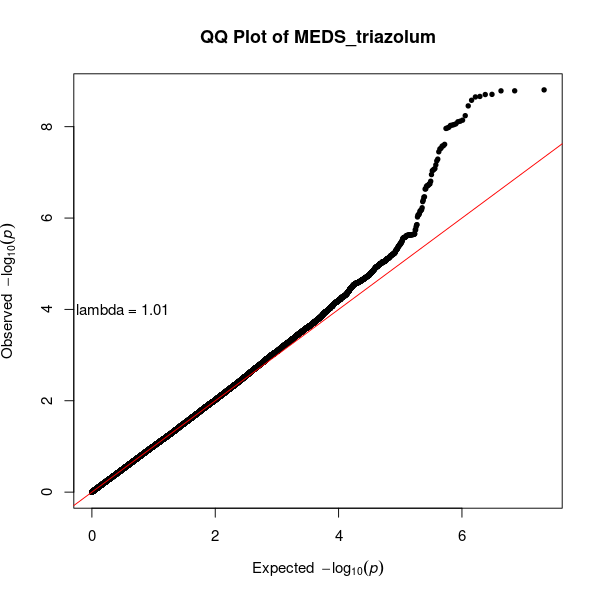

**Figure 1w.** MEDS_potassium_clorazepate

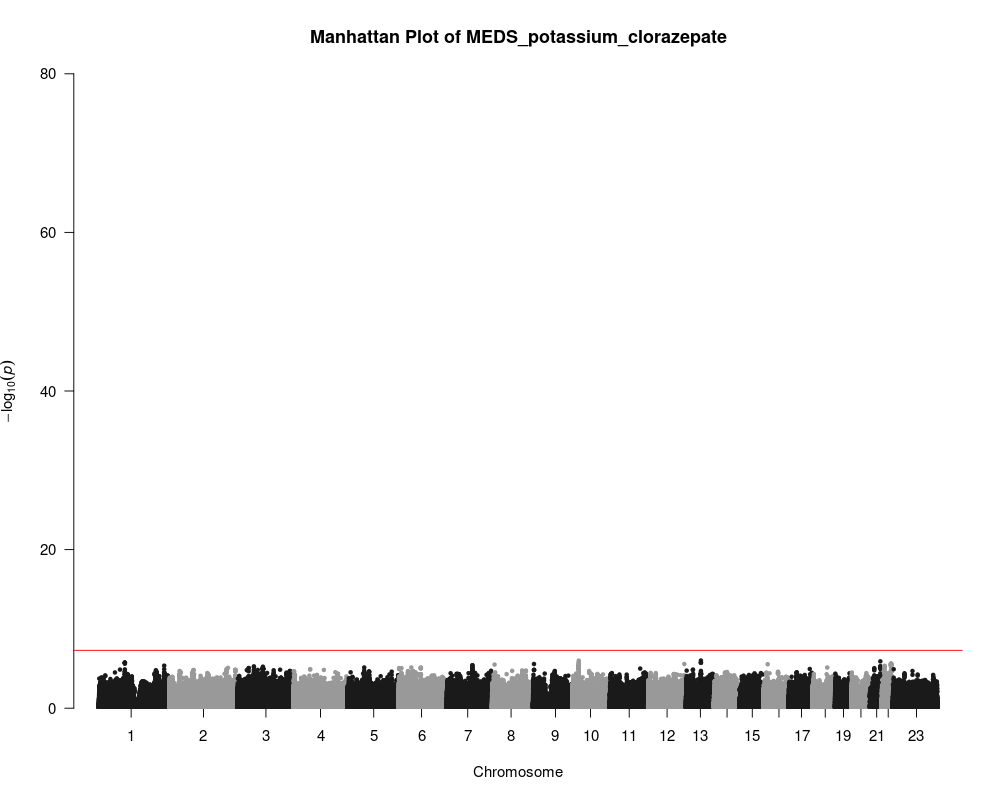

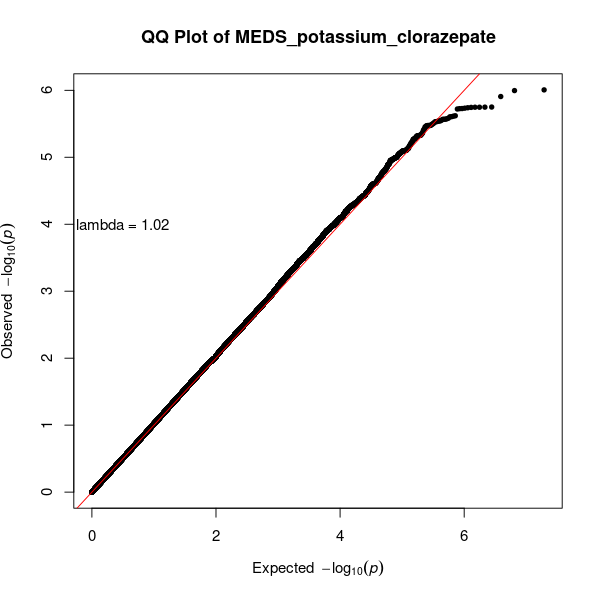

**Figure S2.** Trait heritability estimates

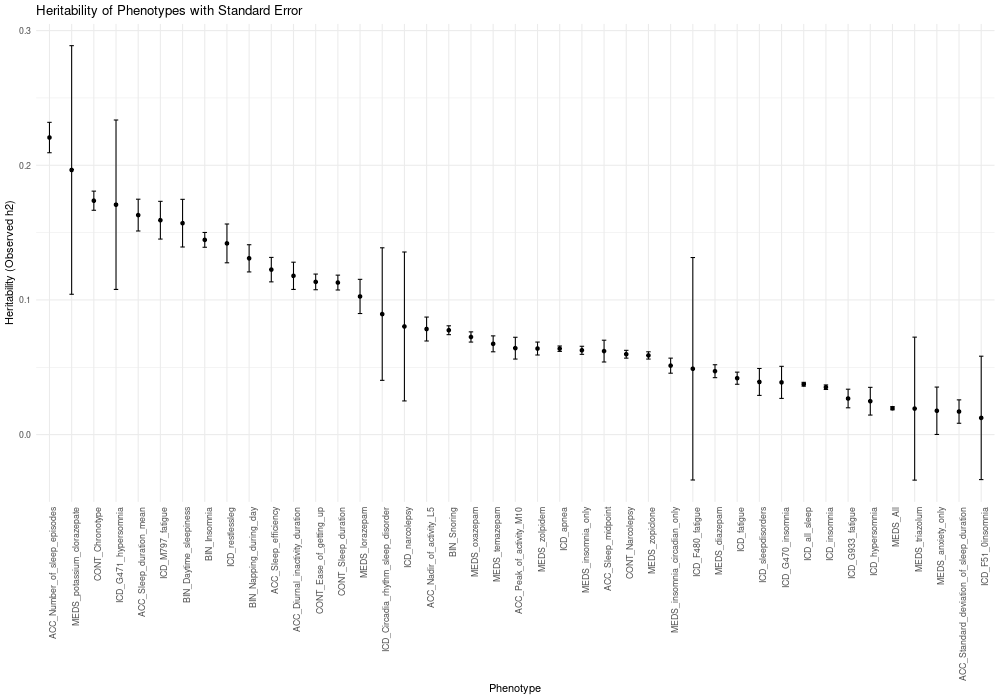

**Figure S3a.** Gene expression enrichment with clinical insomnia.

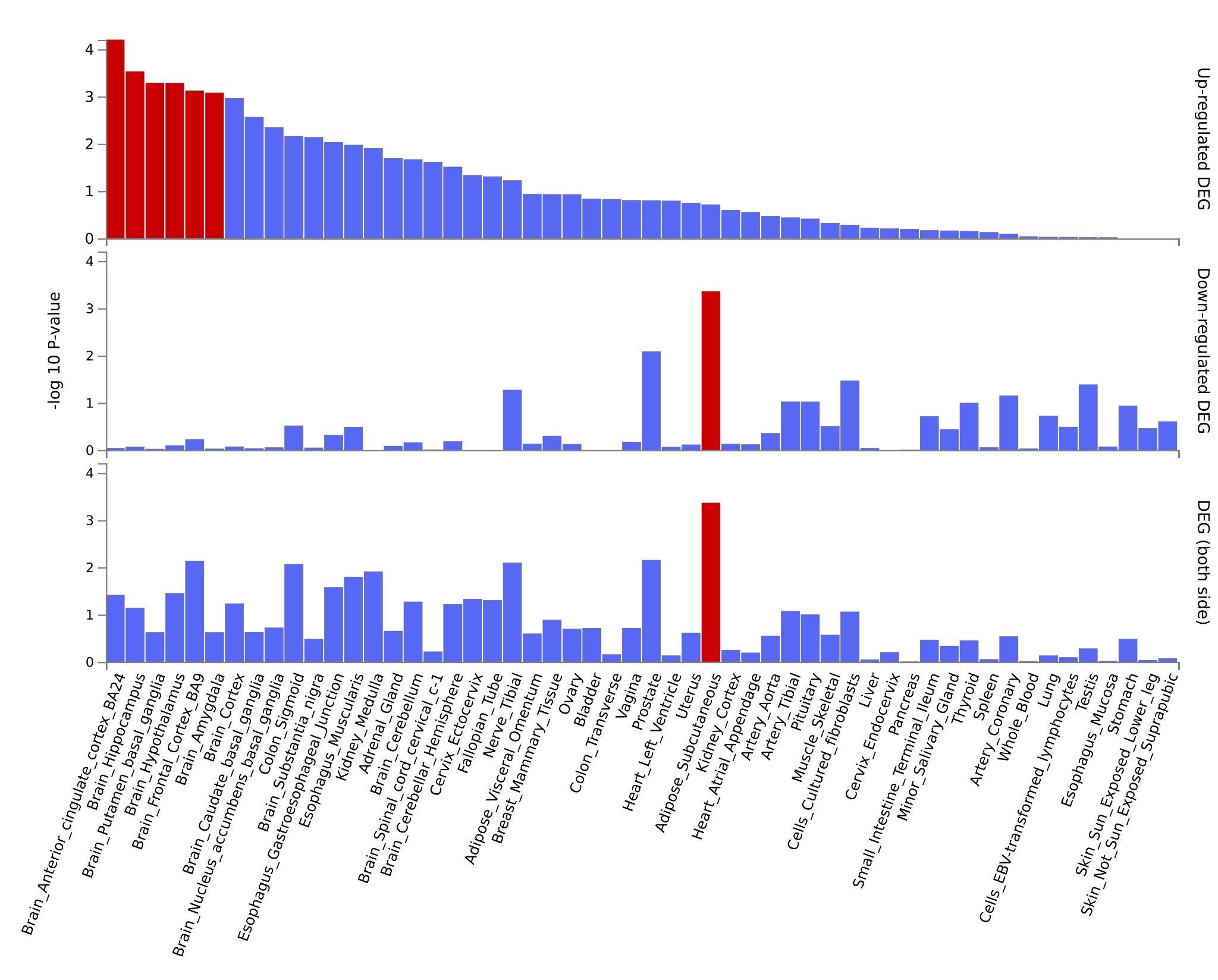

**Figure S3b.** Transcription factor enrichment with clinical insomnia

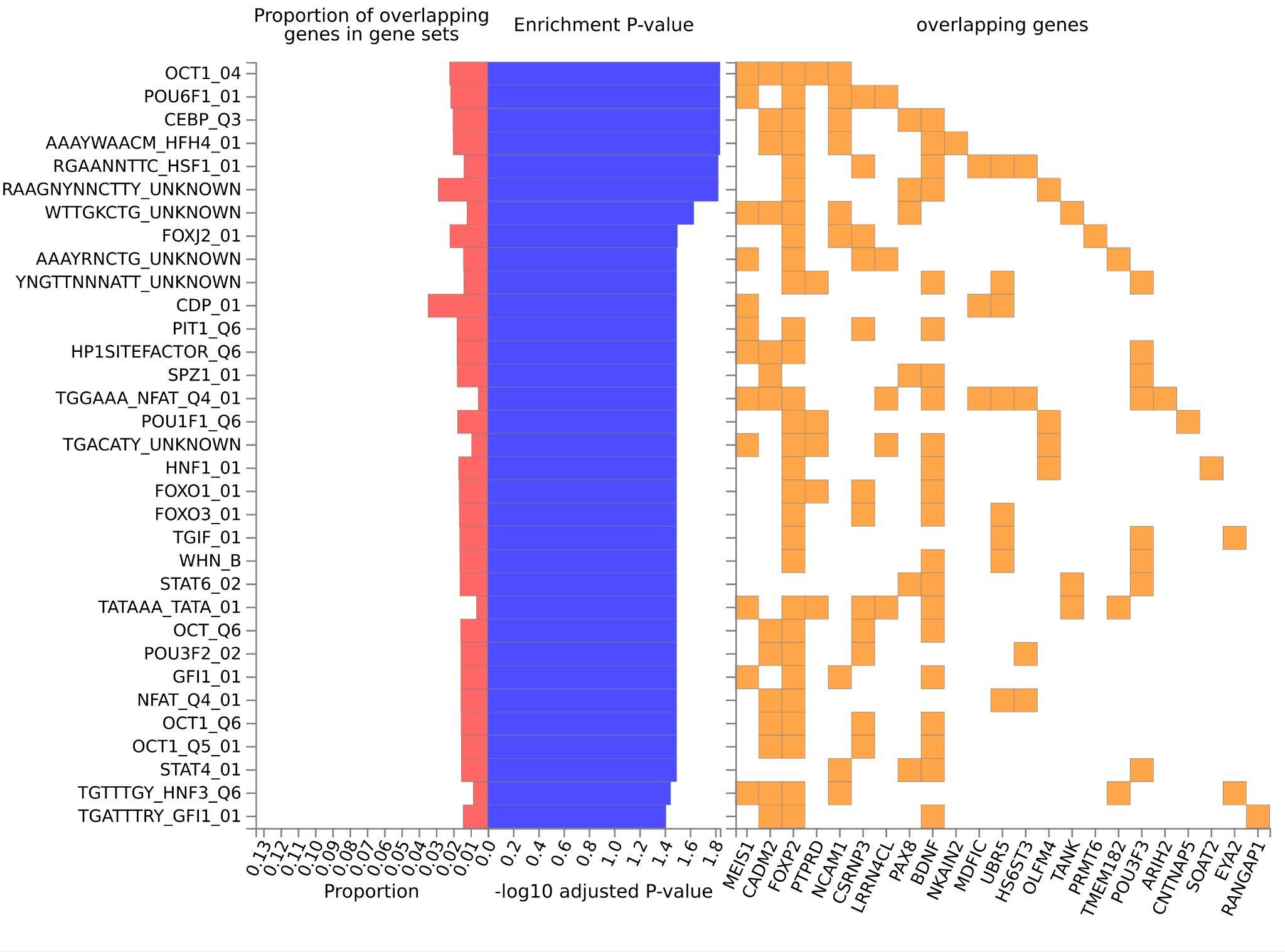

**Figure S3c.** Cell type enrichment in clinical insomnia

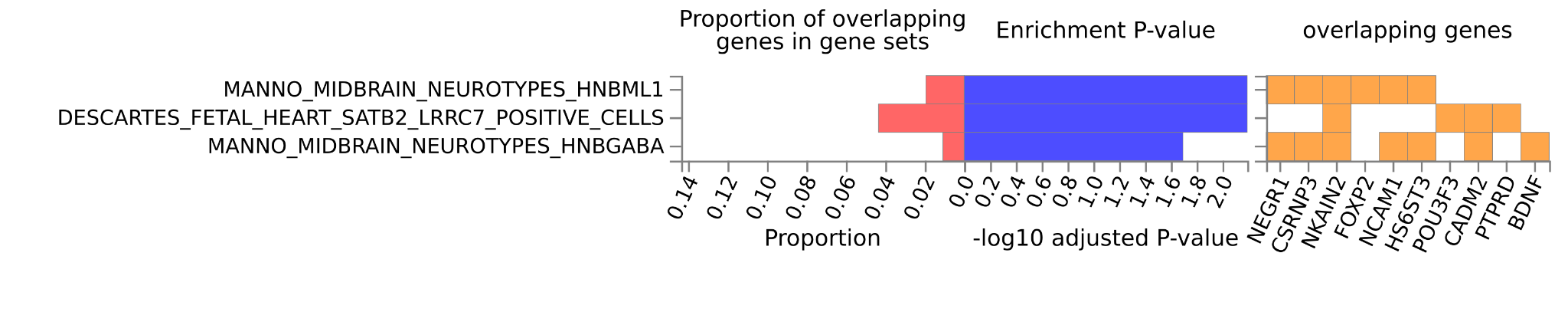

**Figure S4a.** Gene expression enrichment in RLS

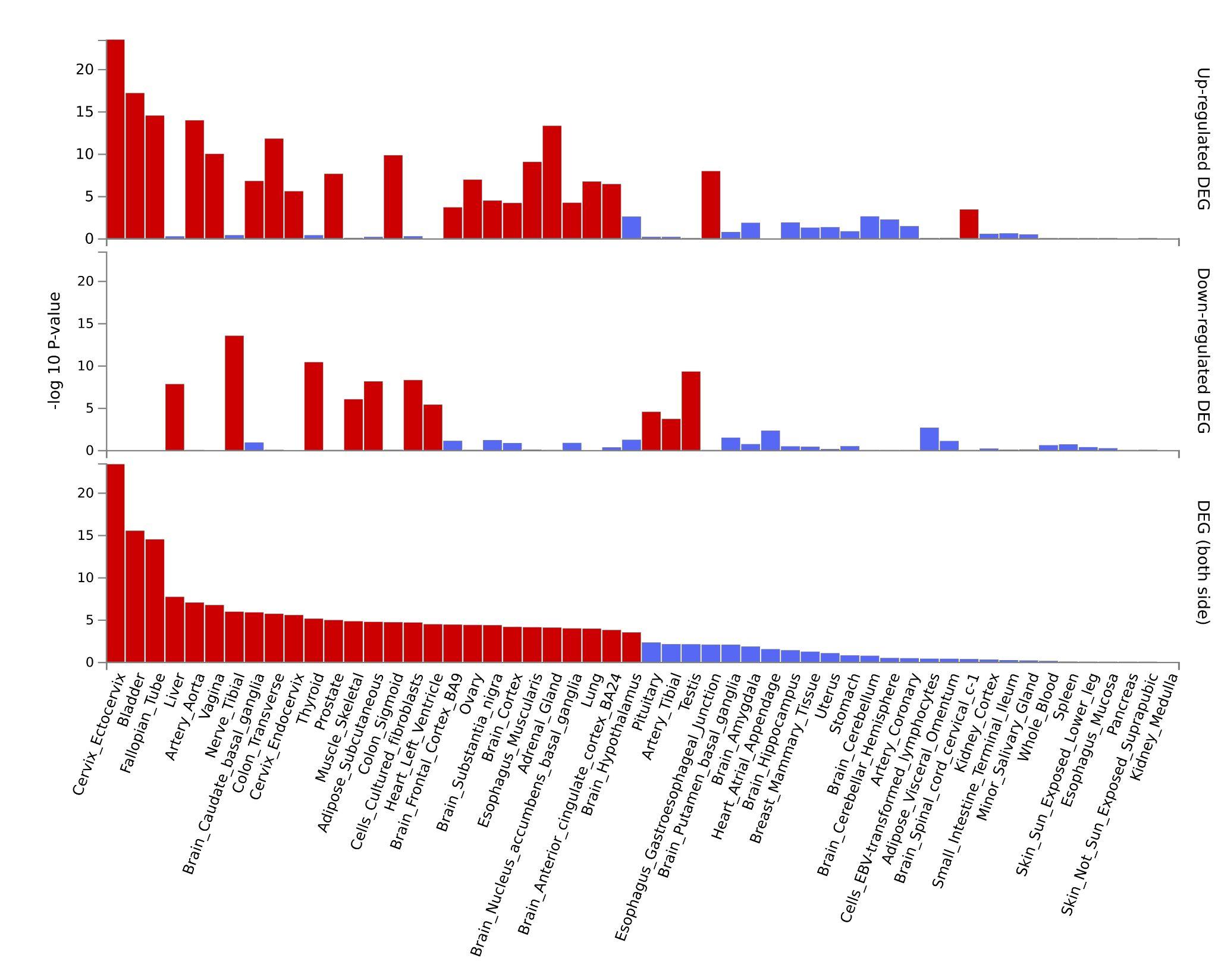

**Figure S4b.** Transcription factor enrichment in RLS

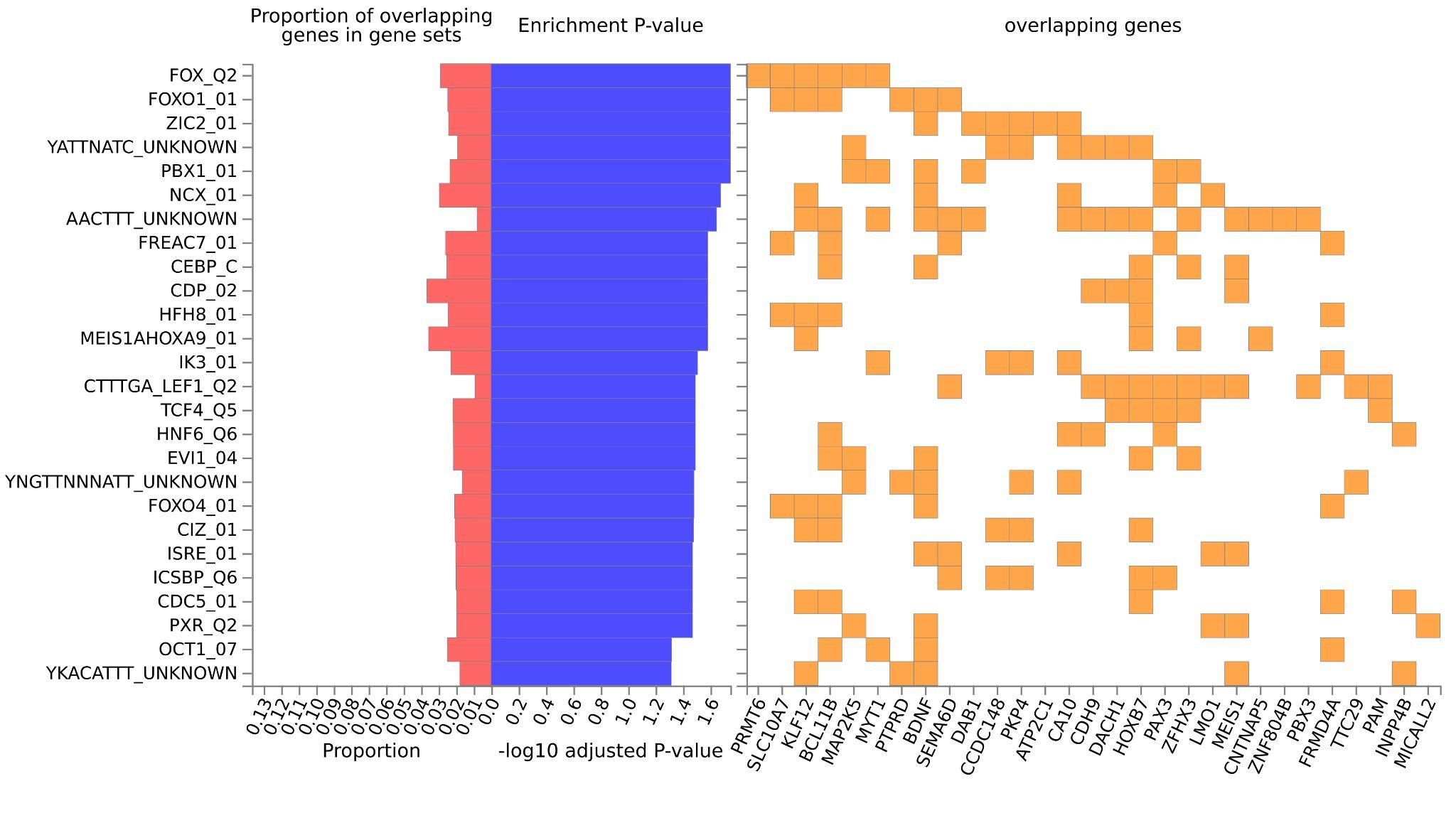

**Figure S4c. GO biological processes in RLS**

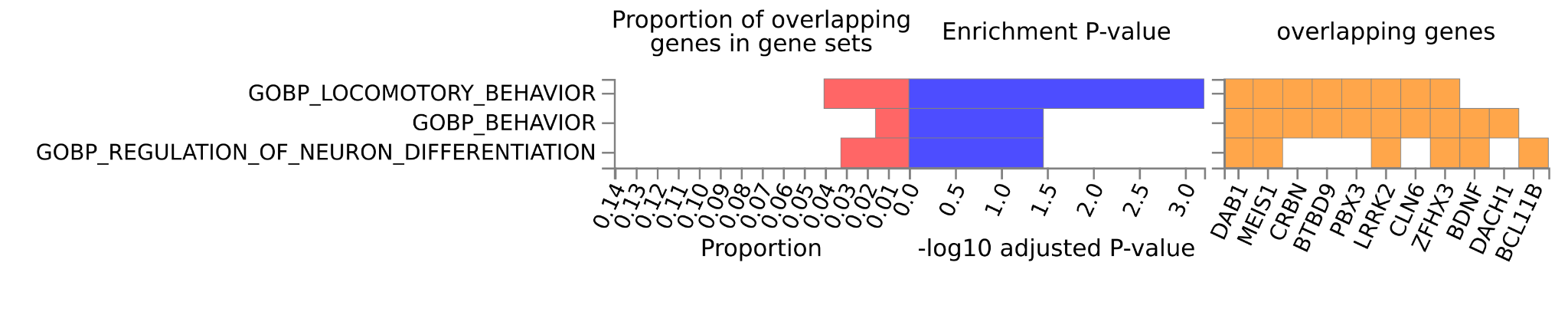

**Figure S4d. Cell type signatures in RLS**

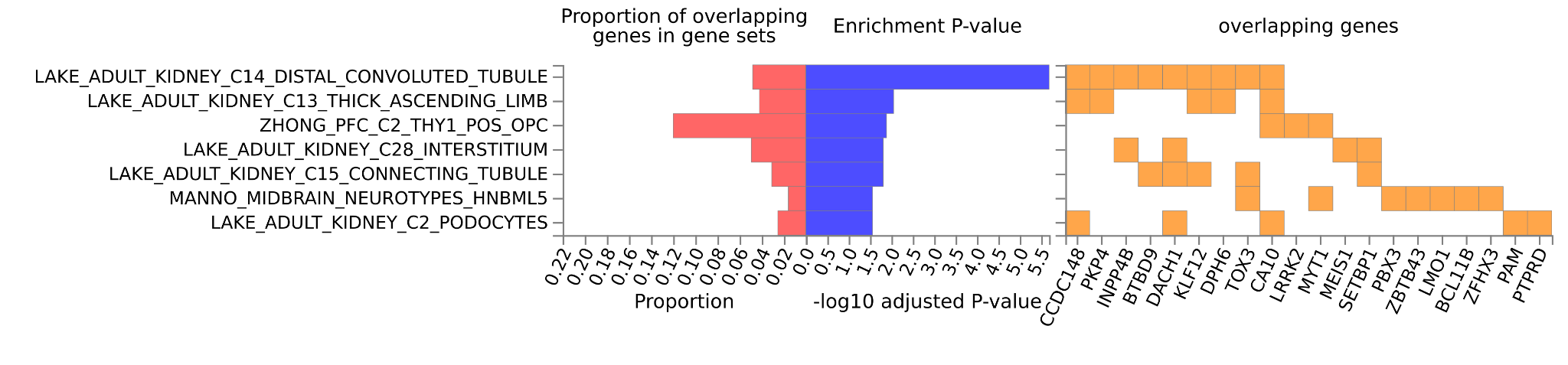

**Figure S5.** Gene expression enrichment with sleep apnea

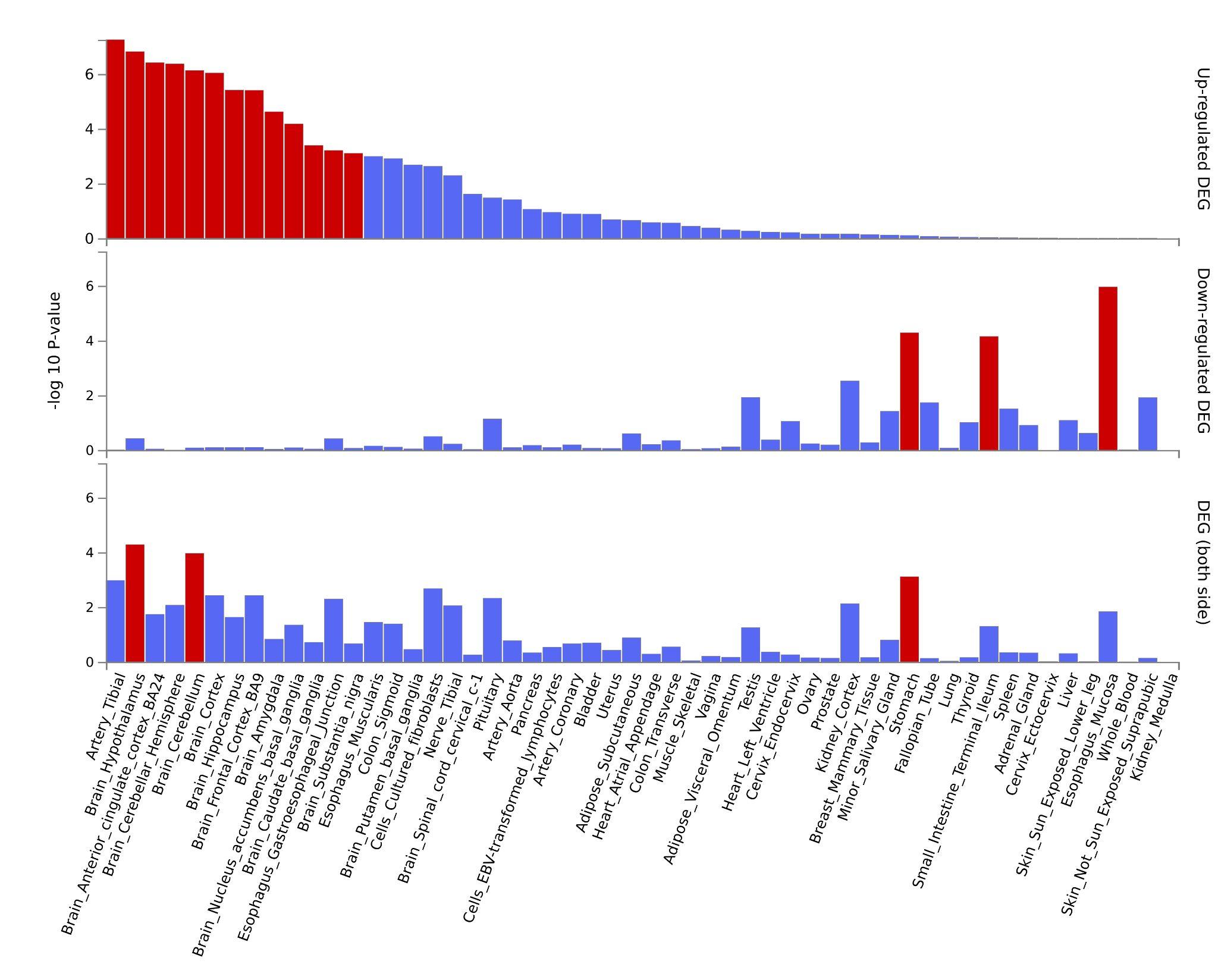

**Figure S6.** *Genetic correlation between sleep disorders and selected neuropsychiatric traits.*

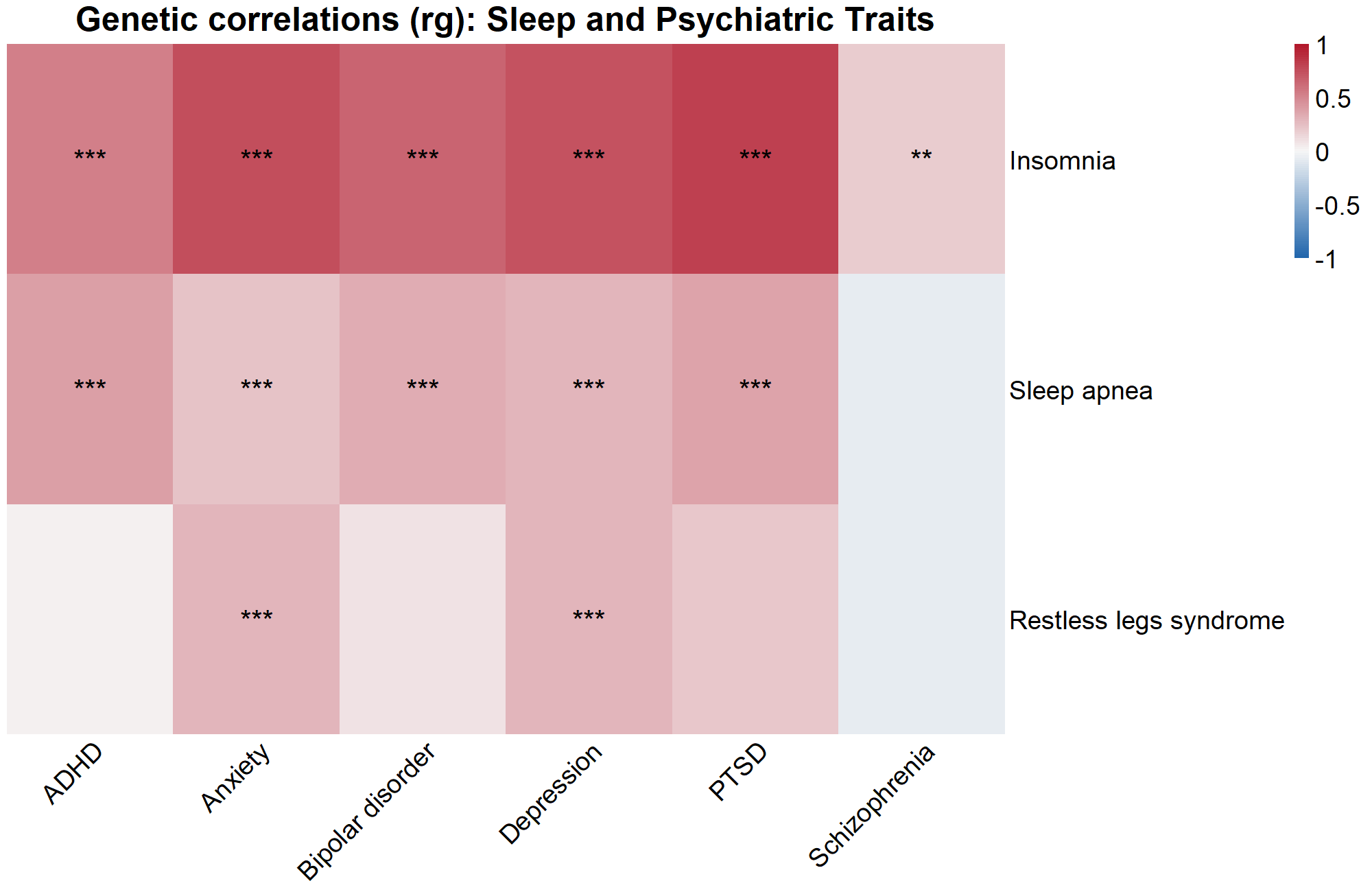

***Figure S7. Estimating environmental factors in sleep disorders with Mendelian randomization.*** ***A)*** *Low ferritin increases the risk for RLS.* ***B)*** *Low ferritin does not increase the risk for insomnia.* ***C)*** *High BMI (influenced by both environment and genetics) increases the risk for sleep apnea. There was no evidence of pleiotropy in any of the tested causal relationships (P MR Egger intercept = ns.)*

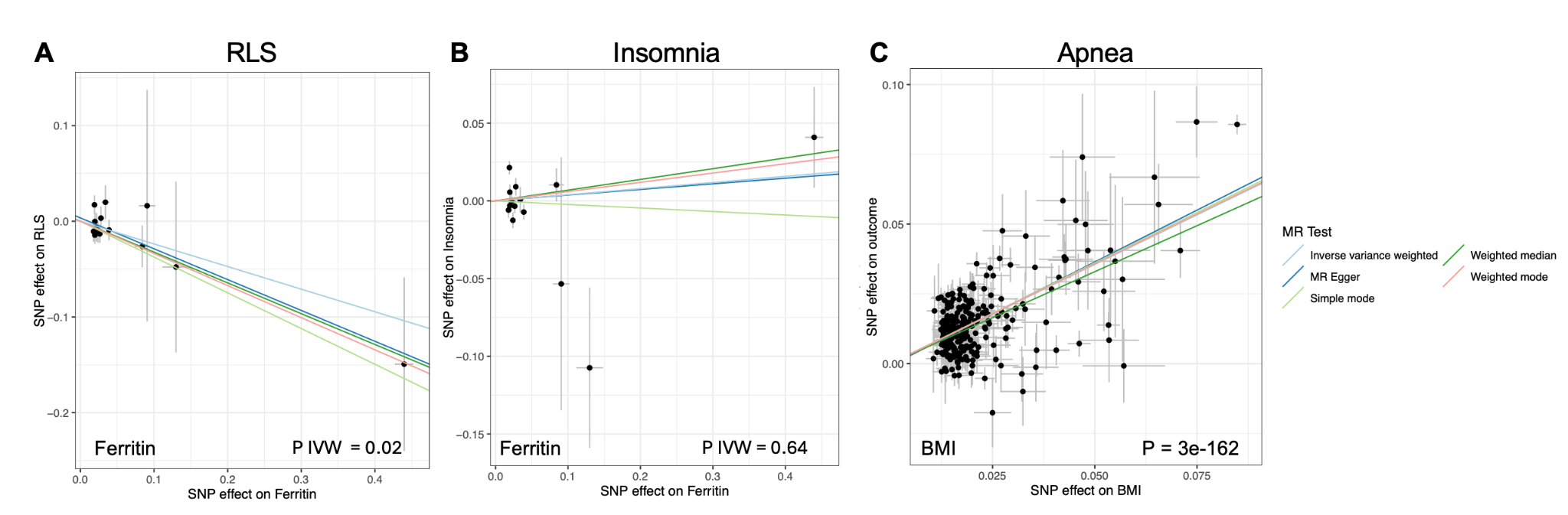
